# Genome-Wide Association Analysis of Tic Disorders Reveals 6 Independent Risk Loci and Highlights Tic-Associated Cell Types and Brain Circuitry

**DOI:** 10.64898/2026.04.09.26350245

**Authors:** Dongmei Yu, Nora I. Strom, Zachary F. Gerring, Apostolia Topaloudi, Matthew W. Halvorsen, Sudhanshu Shekhar, Tyne W. Miller-Fleming, Miao Tang, Luz M. Porras, Franjo Ivankovic, Behrang Mahjani, Teemu Palviainen, Elizabeth C. Corfield, Christos Androutsos, Alan Apter, Helga Ask, Valentina Baglioni, Juliane Ball, Cathy L. Barr, Csaba Barta, Entela Basha, James R. Batterson, Noa Benaroya-Milshtein, Fortu Benarroch, Dorret I. Boomsma, Anders D. Børglum, Cathy L. Budman, Jan K. Buitelaar, Judith Buse, Jonas Bybjerg-Grauholm, Francesco Cardona, Danielle C. Cath, Larisa H. Cavallari, Keun-Ah Cheon, Barbara J. Coffey, Niklas Dahl, Christel Depienne, Andrea Dietrich, Laura Domènech, Petros Drineas, Gudmundur Einarsson, Erik M. Elster, Siyan Fan, Dana Feldman, Thomas V. Fernandez, Jakub P. Fichna, Natalie J. Forde, Abel Fothi, Carolin Fremer, Emily Gantz, Blanca Garcia-Delgar, Marianthi Georgitsi, Donald L. Gilbert, Jeffrey Glennon, Danea Glover, Marco A. Grados, Erica L. Greenberg, Jakob Grove, Daniel F. Gudbjartsson, Andreas Hartmann, Alexandra Havdahl, Tammy Hedderly, Bastian Hengerer, Luis Diego Herrera-Amighetti, Isobel Heyman, Pieter J. Hoekstra, Hyun Ju Hong, Alden Y. Huang, Chaim Huijser, David Isaacs, Piotr Janik, Joseph Jankovic, Cathrine Jespersgaard, Seulgi Jung, Ahmad Seif Kanaan, Mira Kapisyzi, Christina Kappler-Friedrichs, Jaakko Kaprio, Iordanis Karagiannidis, Najah Khalifa, Young Shin Kim, Robert A. King, Nadine Kirchen, Carolin Sophie Klages, Yun-Joo Koh, Anastasia Koumoula, Samuel Kuperman, James F. Leckman, Bennett L. Leventhal, Holan Liang, Christine Lochner, Maria Loreta Lopez, Marcos Madruga-Garrido, Irene A. Malaty, Osman Malik, William M. McMahon, Sandra Melanie Meier, Marieke D Messchendorp, Dararat Mingbunjerdsuk, Pablo Mir, Astrid Morer, Norbert Müller, Kirsten Müller-Vahl, Alexander Münchau, Laura Muñoz-Delgado, Tara Murphy, Peter Nagy, Benjamin M. Neale, Erika L. Nurmi, Kevin Sean O’Connell, Michael S. Okun, Shanmukha S. Padmanabhuni, David L. Pauls, Kerstin Plessen, Geert Poelmans, Cesare Porcelli, Danielle Posthuma, Petra J. W. Pouwels, Joanna Puchala, Renata Rizzo, Mary Robertson, Veit Roessner, Alyssa Rosen, Guy Rouleau, Sven Sandin, Paul Sandor, Martin Scheiene, Simon Schmitt, Joshua A. Senior, Harvey S. Singer, Jordan W. Smoller, Sara Sopena, Tamar Steinberg, Natalia Szejko, Urszula Szymanska, Zsanett Tarnok, Olafur Thorarensen, Meitar Timmor, Jay A. Tischfield, Zeynep Tümer, Anne Uhlmann, Odile A. van den Heuvel, Ysbrand D. van der Werf, Marta Correa Vela, Dick J. Veltman, Ana Vigil-Pérez, G. Bragi Walters, Belinda Wang, Thomas Werge, Joanna Widomska, Tomasz Wolanczyk, Yulia Worbe, Jinchuan Xing, Jin Yin, Cezary Zekanowski, Nuno R. Zilhäo, Samuel H. Zinner, Joseph D. Buxbaum, Dorothy E. Grice, Christina Hultman, Hreinn Stefansson, Ole Andreassen, Aarno Palotie, Gary A. Heiman, Michael J. Gandal, Lea K. Davis, Paola Giusti-Rodríguez, Manuel Mattheisen, James J. Crowley, Peristera Paschou, Jeremiah M. Scharf, Carol A. Mathews

## Abstract

Tourette Syndrome and other tic disorders (TD) are common, highly heritable neurodevelopmental conditions with complex genetic architectures. We conducted a genome-wide association study of 13,247 TD cases and 536,217 European ancestry controls and identified six independent genome-wide significant loci, including a pleiotropic signal at 3p21 shared with attention-deficit/hyperactivity disorder, among other traits. Gene prioritization highlighted 20 genes, including *PCDH9, HCN1, NCKIPSD, WDR6, DALRD3*, and *CELSR3*. Integrative analyses provide genetic support for the role of cortico-striato-thalamo-cortical circuits in TD pathophysiology and further localize TD genetic risk to specific cell types, including dopamine D1- and D2-receptor-positive medium spiny neurons, cortical pyramidal neurons, and oligodendrocyte-lineage cells. We further demonstrate extensive genetic correlations with neurodevelopmental and psychiatric traits, but not with neurological disorders. These findings advance our understanding of the genetic basis of TD, pinpointing specific genes and cell types that drive pathophysiology and providing a foundation for future mechanistic studies.

## Introduction

Tic disorders (TDs) are a spectrum of heritable neurodevelopmental disorders characterized by the presence of motor and/or vocal tics that typically begin in childhood and have shared genetic risk^1–3^. Persistent TDs are defined as those with tic symptoms lasting for at least a year, and include persistent motor or vocal TD (PMVTD; consisting of either motor or vocal tics) and Tourette Syndrome (TS; consisting of both motor and vocal tics)^4–6^. Other TDs in either the DSM or the ICD diagnostic schemas include transient/provisional TD (TTD, F95.0), TD not otherwise specified (F95.9), and other tic disorders (F95.8). TS typically begins in early childhood and affects approximately 0.6–1% of the global population ^4,5^, while PMVTD affects an additional 1-1.5% ^7–9^. TDs can cause significant functional disability^6^, and are highly comorbid with obsessive-compulsive disorder (OCD), attention-deficit/hyperactivity disorder (ADHD), autism spectrum disorder (ASD), and anxiety disorders^6,10–12^, which further contribute to impairment^4,10,11^. Persistent TDs have among the highest estimated heritability of non-Mendelian neurodevelopmental disorders (60-77%)^13–16^, whereas heritability estimates for TTD range from 25-37% ^17^. Genome-wide association studies (GWAS) have provided SNP-based heritability estimates for TD ranging from 14% to 58%^1,18–20^, suggesting that common genetic variants contribute significantly to TD risk. However, the genetic underpinnings of TD remain elusive, in part due to their complex polygenic architecture.

Neuroimaging, postmortem, and animal studies consistently highlight cortico-striatal-thalamo-cortical (CSTC) circuitry as central to the pathophysiology of TD and related disorders, particularly OCD^21^ and ADHD^22^. Structural and functional magnetic resonance imaging studies (sMRI and fMRI, respectively) in individuals with TD reveal differences in the striatum, thalamus, and prefrontal cortex; these regions are critical for motor learning^23^, habit formation, cognitive control, and emotion regulation^24^. Altered striatal volume in structural neuroimaging studies of TS and disrupted connectivity between striatum and sensorimotor cortex in fMRI studies are consistent with the sensorimotor gating impairment observed in this population^25–27^, which has been implicated in tic genesis^28^. Thalamic hyperactivity and atypical prefrontal engagement identified in TS imaging studies align with observed deficits in cognitive control, and changes in the orbitofrontal cortex, cingulate cortex, and ventral striatum correspond with abnormalities in reward processing, habit formation, and emotion regulation, all of which are also implicated in OCD and ADHD^24,29–32^. Further underscoring the critical role of CSTC circuits in TD pathophysiology, resting-state fMRI studies demonstrate increased local connectivity and reduced global coordination within these circuits^33^. Bridging genetic findings with hypotheses generated from neuroimaging data can advance insights into the contribution of genetic variants to circuit-level dysfunction in TD.

GWAS have significantly advanced our understanding of TD genetics, providing insights into their genetic architecture, and identifying potential genetic susceptibility loci^1,18^. However, early GWAS were underpowered, and initial genome-wide significant loci have yet to achieve consistent genome-wide replication^1,19,20^. Gene-based analyses in prior GWAS also identified additional genes of interest involved in neuronal development and synaptic plasticity; however, these also have yet to be replicated^1,19,20^. Despite the lack of robustly replicated loci, these studies provide important preliminary data, not only with respect to potential genes that may be involved in TD pathogenesis, but also into the role of putative TD genes in the neural circuitry and biological mechanisms underlying TD^1,34,35^.

In this study, we present a TD GWAS that includes 13,247 cases and 536,217 controls, substantially increasing statistical power to identify new genome-wide significant TD risk loci. We also present comprehensive post-GWAS analyses, including the prioritization of TD risk genes through positional and functional gene mapping, identification of specific mouse and human brain cell types whose transcriptomes are enriched with TD polygenic signals, and converging biological evidence linking TD-associated genetic variation to neurodevelopmental pathways and CSTC circuitry. By integrating gene-based, transcriptomic, and single-cell-based analyses with genome-wide association studies, we bridge the gap between genetic associations and biological pathways, offering novel insights into TD pathophysiology.

## Results

### TD GWAS identifies six novel risk loci

We conducted a GWAS meta-analysis comprising 7,080,972 SNPs in 13,247 individuals with TD and 536,217 ancestry-matched controls. This study integrated 14 European-ancestry case-control cohorts, including 8 new cohorts and 6 previously published GWAS datasets^1,19,20,36^ (Supplementary Notes, Supplementary Table 1). Most cases (93%, N = 12,343) were diagnosed with TS or PMVTD, with the remainder (7%, N = 904) having other TD diagnoses. No significant heterogeneity was observed across cohorts based on Cochran’s Q-test and *I*^*2*^ statistic (Supplementary Fig. 1) despite varied ascertainment methods (Supplementary Table 1, Supplementary Notes). Univariate Gaussian mixture modeling in MiXeR^37^ estimated that approximately 12,131 causal SNPs underlie 90% of the SNP-based heritability of TD.

To evaluate the impact of phenotypic heterogeneity, we performed stratified meta-analyses based on ascertainment method (semi-structured interviews vs. national registries) (Supplementary Table 2) and case inclusion criteria: (i) strictly defined TS (subset TS), (ii) any tic disorder (subset TD), and (iii) any tic disorder ascertained in the context of another psychiatric disorder (subset TD_comorbid) (Supplementary Table 3). All pairwise analyses demonstrated high genetic correlations, with *r*_*g*_ = 0.88 between semi-structured interviews and national registries (*P* = 1.4 × 10^-28^), 0.78 between subset TS and TD (*P* = 2.1 × 10^-11^), 0.88 between subset TS and TD_comorbid (*P* = 5.3 × 10^-19^), and 0.99 between subset TD and TD_comorbid (*P* = 7.9 × 10^-9^), indicating a largely homogenous genetic architecture across different TD definitions (Supplementary Notes, Supplementary Table 4). Confirmatory factor analysis (CFA) of the three case inclusion stratified subsets using Genomic SEM^38^ under a one-factor model revealed strong and highly significant loadings on a common latent factor (standardized loadings = 0.85–0.88, all P < 1.0 × 10^−13^; Supplementary Table 5), suggesting that the majority of genetic variance was captured by an underlying common dimension. Residual variances were not significant (standardized residuals = 0.22– 0.28, all P > 0.1), indicating limited evidence for subset-specific genetic effects.

We identified six independent novel genome-wide significant loci across 37 SNPs that exceeded the genome-wide significance threshold (p < 5.0 × 10^-8^) (Fig. 1, Table 1, Supplementary Table 6). Conditional and joint multiple-SNP analysis^39^ (GCTA-COJO) confirmed the independence of these six loci but did not reveal additional independent signals (Supplementary Table 7). Regional and forest plots are presented in Supplementary Figs. 2-8.

**Fig. 1.**
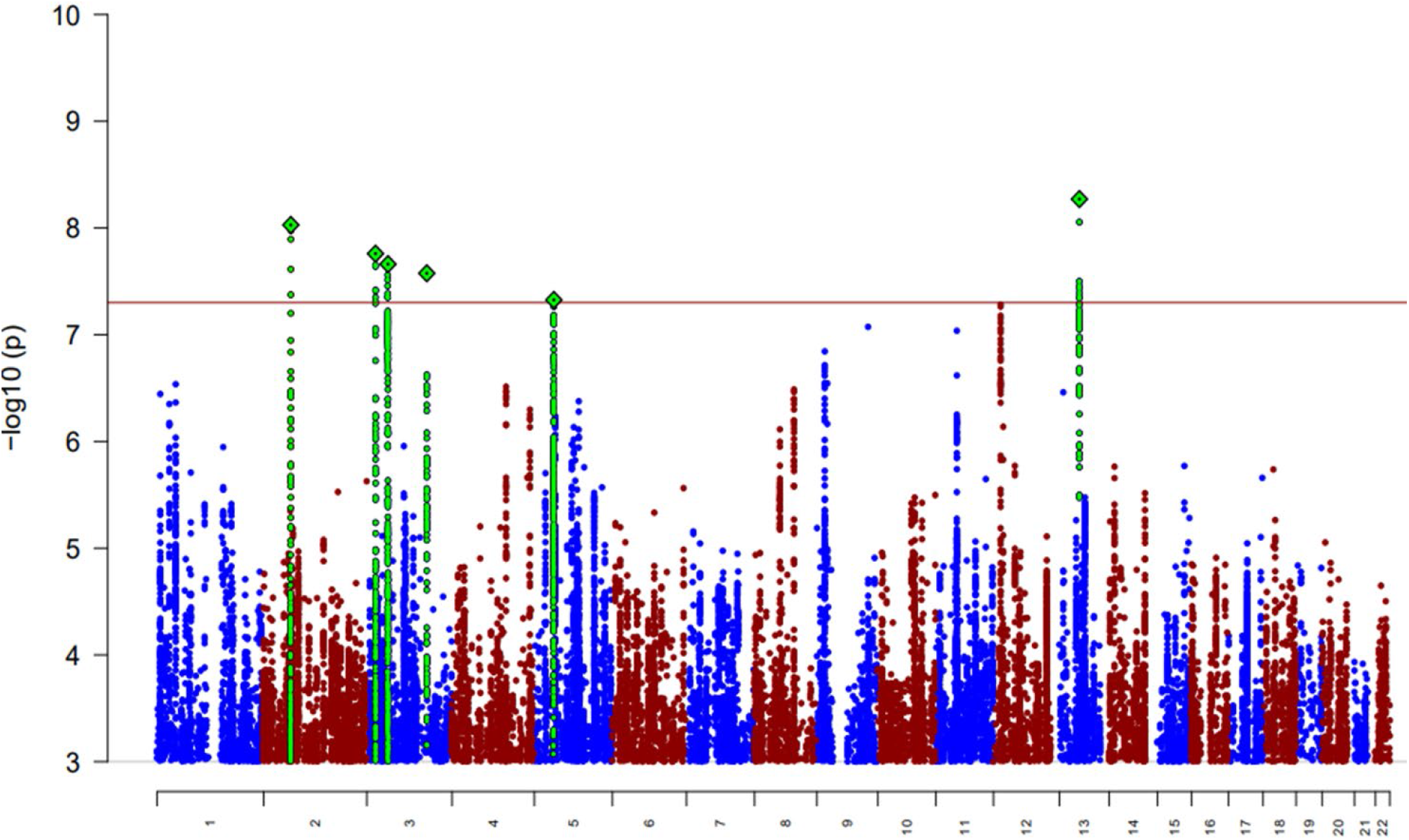
Manhattan plot for GWAS meta-analysis of 14 cohorts including 13,247 cases and 536,217 controls in total. The *y-axis* represents −log10(association *P*-value) from the meta-analysis using an inverse variance-weighted fixed-effects model. The green diamonds represent the index SNPs in each of the genome-wide significant loci, and the green dots represent the SNPs in LD (*r*^*2*^ > 0.5) with the index SNPs. The red horizontal line represents the threshold for genome-wide significant association (*P* = 5.0 × 10^−8^). Only the SNPs with association P-value < 0.001 were plotted.

**Table 1.**
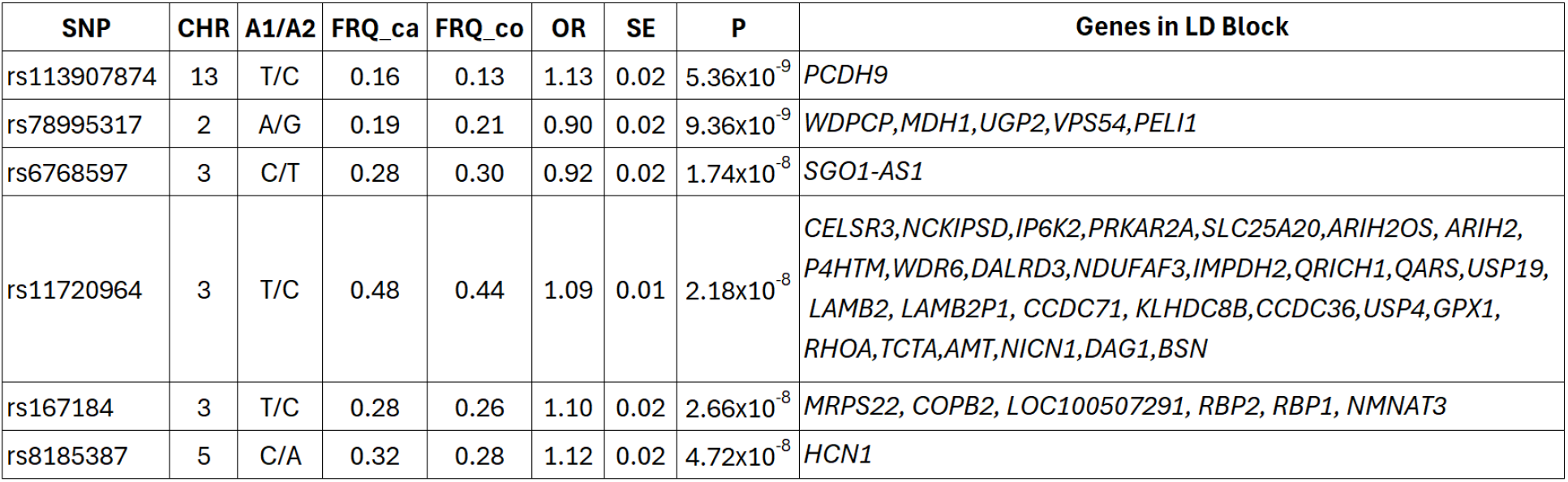
Genome-wide-significant loci associated with TD. SNP: The index SNP with the lowest P-value within the significant locus. CHR: Chromosome where the SNP is located. A1/A2: Effect allele (A1) and alternative allele (A2). FRQ_ca: Frequency of the effect allele (A1) in cases. FRQ_co: Frequency of the effect allele (A1) in controls. OR: Odds Ratio of the effect allele (A1). SE: Standard error of the effect size (i.e., ln(OR)). P: Associated P-value. Genes in LD Block: Genes overlapping the LD block, including 5kb flanking regions on both ends.

The most significant locus was on 13q21 (rs113907874, *P* = 5.4 × 10^−9^), located within exon 4 and intron 4 of *PCDH9*, a protocadherin gene implicated in synaptic signaling^40^. The second most significant locus was on 2p14 (rs78995317, *P* = 9.4 × 10^−9^), within a linkage disequilibrium (LD) block encompassing *UGP2* and *VPS54*, genes associated with developmental and epileptic encephalopathy^41^ and progressive motor neuron degeneration^42^, respectively. Three independent loci on chromosome 3 reached genome-wide significance: 3p24 (rs6768597, *P* = 1.7 × 10^−8^), which overlaps *SGO1-AS1* (previously associated with ADHD^43,44^, educational attainment^45^, smoking initiation^46^, and insomnia^47^); 3q23 (rs167184, *P* = 2.7 × 10^−8^), situated near genes involved in neurodevelopment and intracellular transport^48–50^ (*MRPS22, COPB2, RBP1*, and *RBP2*), and 3p21 (rs11720964, *P* = 2.2 × 10^−8^). Notably, the 3p21 signal falls within a gene-dense region, harboring 28 protein-coding genes within a complex chromatin architecture (Supplementary Fig. 6), and encompassing *CELSR3*, which was previously implicated in TS by a rare-variant WES study^51^. This locus has been associated previously with multiple psychiatric and cognitive traits, including TS^52–54^, OCD^55^, major depressive disorder^56^ (MDD), anxiety disorders^57^, substance use disorders^58,59^, insomnia^47^, general cognitive function^60^, and cerebellar volume^61^. Finally, a sixth novel locus, on 5p11-p12 (rs8185387, *P* = 4.7 × 10^−8^), encompasses *HCN1*, which regulates neuronal excitability and brain development and has been implicated in schizophrenia^62^, depression and anxiety^63^, epilepsy^64^, and ASD^65^.

Beyond these findings, we assessed other previously reported TD-associated loci (Supplementary Tables 8-9). We observed an association, though not genome-wide significant, for the previously reported TD risk SNP rs79244681 (*MCHR2-AS1, P* = 5.7 × 10^-5^)^20^. Several other loci, including *FLT3* (rs2504235, *P* = 0.016), *WWC1* (rs6555807, *P* = 1.2 × 10^-3^), *NIPBL* (rs80062984, *P* = 5.0 × 10^-3^), and *FN1* (rs17519171, *P* = 1.9 × 10^-4^), showed nominal evidence of association in this GWAS, while two previously reported TS loci (*COL27A1* and *DRAM1*) did not replicate.

We applied PAINTOR^66^ to prioritize candidate causal variants at the six genome-wide significant loci. Four loci yielded high-confidence variants with posterior probabilities (PP) > 0.9, including rs113907874 at 13q21 (PP = 0.92), rs9854943 at 3p24 (PP = 0.99), rs4858798 at 3p21 (PP = 0.97), and rs9878661 at 3q23 (PP = 1.00), suggesting that these are strong candidates for functional follow-up (Supplementary Tables 10-11). No variants at 2p14 or 5p11-p12 exceeded the causality threshold (PP > 0.5), possibly reflecting limited power or insufficient functional annotation at these loci.

### SNP-heritability and Polygenic Risk

We estimated the SNP-heritability using LD Score Regression^67^ (LDSC) for the overall and stratified meta-analyses. The SNP-heritability of the overall meta-analysis on the liability scale was 14.9% (SE = 1.0%), similar to the *h*^*2*^_*SNP*_ estimated from a previous TD GWAS^20^ (13.8%), but lower than the previously reported h^2^ in a TS-only GWAS^1^ (21%). Stratified analyses demonstrated higher SNP-heritability estimates for the TS-only subset (h^2^ _SNP_ = 22.2%, SE = 2.1%) compared with the TD subset (h^2^_SNP_ = 9.9%, SE = 1.9%) and TD_comorbid subset (h^2^_SNP_ = 17.7%, SE = 2.5%), suggesting that ascertainment and case definition influence SNP-heritability estimates.

Polygenic Score (PGS) analyses were conducted in four cohorts from the TS subset with at least 500 cases (Supplementary Table 12). We utilized Leave-One-Out (LOO) meta-analyses as the discovery samples, and adjusted SNP effect sizes via PRScs^68^ using HRC European samples as the reference. In all cohorts, the adjusted PGS, derived by controlling for population stratification and standardization, was significantly associated with TD diagnosis, with odds ratios ranging from 1.41 to 1.48, explaining 3.2%-3.9% of the phenotypic variation. Individuals in the top PGS decile had a 2.6 to 3.5-fold increased risk of TD compared to those in the bottom decile (Supplementary Fig. 9).

### Gene-based Analyses and Gene Prioritization

To identify genes contributing to TD risk, we combined two positional gene mapping approaches with five functional mapping approaches (Supplementary Table 13). The positional gene mapping approaches included MAGMA^69^ (Supplementary Table 14) and mBAT-combo^70^ (Supplementary Table 15). Functional gene mapping approaches incorporated eQTL and chromatin interaction data through: 1) E-MAGMA^71^ (Supplementary Table 16), which integrated brain-specific eQTL data from the Genotype– Tissue Expression (GTEx) project^72^; 2) Transcriptome-Wide Association Study (TWAS) using Joint Tissue Imputation (TWAS_JTI), utilizing 49 GTEx v8 tissues via a multi-tissue Aggregated Cauchy Association Test (ACAT) (Supplementary Table 17); 3) TWAS+COLOC, which performed TWAS using FUSION^73^ with PsychENCODE brain gene expression data, followed by colocalization analyses using the COLOC R package^74,75^ (Supplementary Table 18); 4) Summary data-based Mendelian Randomization (SMR) analysis^76^ employing eQTL data from eQTLGen^77^ and MetaBrain^78^, followed by heterogeneity in dependent instruments (HEIDI) for robust association testing (SMR+HEIDI, Supplementary Table 19); 5) H-MAGMA^79^ conducted gene-based tests using brain chromatin interaction profiles (Supplementary Table 20).

We identified 194 genes with significant associations with TD (False Discovery Rate (FDR) < 0.05) using positional gene mapping in MAGMA or mBAT-combo, and 218 genes from at least one of the 5 functional mapping approaches (Fig. 2a-2b). From these results, we identified 17 high-confidence TD risk genes supported by at least one positional test and three or more functional tests (Fig. 2c; Supplementary Table 13). This rigorous approach ensured robust support from multiple lines of genomic evidence. This list included 6 out of 28 genes at the pleiotropic 3p21 locus, including *NCKIPSD* (encoding a protein critical for dendritic spine maintenance), *WDR6* (involved in protein complex assembly and signal transduction), and *DALRD3* (implicated in early brain development). From the other loci identified through positional and functional mapping, 7 of the 11 prioritized genes are related to neurodevelopment or neurotransmission, including *FOLH1*^80^ (involved in folate metabolism and glutamate regulation) and *PTPRJ*^81^ (involved in cell signaling pathways, axonal guidance, and formation of neural circuits) at 11p11, *RERE* at 1p36^82^ (associated with neurodevelopmental disorders), *EMB* at 5q11^83^ (promoting sprouting of motor nerve terminals at the neuromuscular junction), *PITPNM2* at 12q24^84^ (involved in phospholipid transport and intermembrane lipid transfer in the central nervous system), and *MAPT*^85^ (reduced tau expression levels inhibits neuronal migration and leads to intellectual disability^86^) and *KANSL1*^87^ (associated with developmental delay, intellectual disability, and hypotonia) at 17q21. We further investigated the regulatory landscape of these 17 high-confidence genes by examining independent chromatin interaction profiles from fetal and adult brain tissues, excitatory glutamatergic neurons (EGNs), medium spiny neurons (MSNs), and neural progenitor cells (NPCs) (Fig. 2c). These analyses confirmed enhancer-promoter loops for 12 of the 17 genes, providing mechanistic support for their regulation. The five genes for which loops were not observed in these specific datasets were *EMB, CYSTM1, TMCO6, FOLH1*, and *RBM26*.

**Fig. 2.**
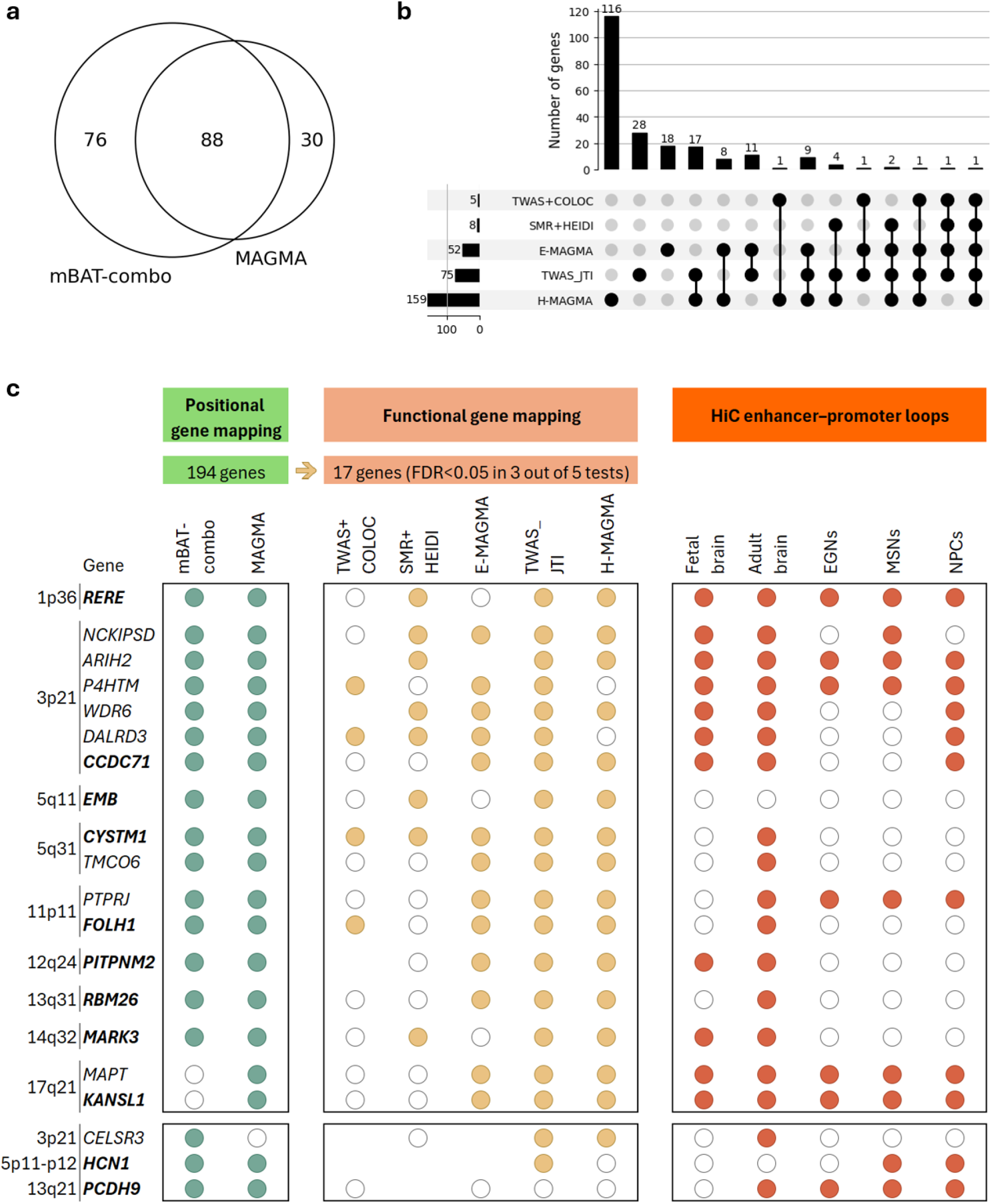
Gene prioritization. a) Positional gene mapping identified a total of 194 genes using two approaches: multivariate Set-Based Association Test (mBAT-combo) and multi-marker analysis of genomic annotation (MAGMA). After false discovery rate (FDR) correction, mBAT-combo and MAGMA identified 164 and 118 genes, respectively. b) Functional gene mapping using five approaches identified a total of 218 genes. These approaches included Transcriptome-Wide Association Study (TWAS) followed by colocalization analysis (TWAS+COLOC) using PsychENCODE, Summary data-based Mendelian Randomization analysis followed by heterogeneity in dependent instruments association test (SMR+HEIDI) using eQTLGen and MetaBrain, E-MAGMA using brain GTEx, TWAS with Joint Tissue Imputation (TWAS_JTI) using GTEx, and H-MAGMA using fetal and adult brain Hi-C data. After FDR correction, these methods identified 5, 8, 52, 75, and 159 significant genes, respectively. c) Summary of prioritized genes. Seventeen high-confidence TD risk genes were prioritized based on significance in at least one positional mapping method (green circles, left panel) and at least 3 functional mapping methods (orange circles, middle panel). Open circles indicate non-significant results for the corresponding gene-mapping test. Blank entries in the middle panel indicate that no eQTL data were available for the gene in the corresponding dataset. The right panel shows evidence of enhancer-promoter interactions (red circles) derived from chromatin conformation (Hi-C) data across 5 brain-relevant contexts: fetal brain, adult brain, excitatory glutamatergic neurons (EGNs), medium spiny neurons (MSNs), and neural progenitor cells (NPCs). Open circles indicate no evidence of enhancer-promoter interactions. Corresponding positional mapping, functional mapping, and Hi-C evidence for 3 additional prioritized genes are shown in the bottom panel, including *CELSR3*, a previously reported TS risk gene in a rare variant study^551^ as well as *HCN1* and *PCDH9*, which are the only genes within the genome-wide significant loci at 5p11-p12 and 13q21, respectively. Genes closest to the index SNP in each locus are highlighted in bold.

In addition to identifying novel high-confidence genes, our gene-based analyses also replicated associations with previously reported TD risk genes. *CELSR3* at 3p21, which plays a critical role in neuronal development, axon guidance, and circuit formation in the brain^88^, was implicated by one positional gene mapping (mBAT-combo) and two functional gene mapping approaches (TWAS-JTI and H-MAGMA). *CELSR3* was previously identified as a potential TS susceptibility gene by Wang et al. based on a *de novo* rare variants study of exome sequencing of TS trios^89^ *HCN1* is the only gene located at the 5p11-p12 locus, and was previously reported as a risk gene for TS from a linkage study^90^ and for early-onset epilepsy^64,91^. *HCN1* was implicated by both positional gene mapping tests and by the TWAS-JTI approach, although the Hi-C analysis did not support this association, possibly attributable to technical limitations of Hi-C in near-centromeric regions or the absence of data from relevant developmental stages or cell types.

### Tissue and cell type enrichment

We next performed tissue and cell-type specificity analyses using both human and mouse brain cell types in MAGMA to explore which tissues and/or cell types showed enrichment of TD genetic risk. We first used single-cell RNA-seq data from the adult mouse central and peripheral nervous systems, encompassing 265 cell types^92^ and observed significant enrichment of TD polygenic risk in 38 of the 265 cell types (FDR < 0.05) (Supplementary Tables 21-23, Supplementary Figs. 10, 11b). Of note, cell types directly involved in the CSTC circuitry (N = 40) were significantly enriched in the 38 TD-associated cell types (N = 16, χ^2^ = 22.85, *P* = 1.7 × 10^-6^). Nine of these cell types correspond to the somatomotor components of the CSTC circuitry (Fig. 3a, highlighted in red), including dopamine D1/D2 receptor-expressing neurons in the dorsal striatum, consistent with the hypothesis that TD is associated with dysregulation of the procedural memory (i.e., motor learning) network in the brain^93^. In addition, a significant proportion of the single-cell RNA-seq data map to non-motor structures and cell types involved in cognitive/associative processes (Fig. 3a, highlighted in blue, N = 4), as well as affective/limbic processes (Fig. 3a, highlighted in orange, N = 12), including dopamine D1/D2 receptor neurons in the ventral striatum. Furthermore, the basal ganglia subregions and cell types enriched with TD polygenic risk included both dopamine D2 Receptor positive (D2R+) MSNs and dopamine D1 Receptor positive (D1R+) MSNs.

**Fig. 3.**
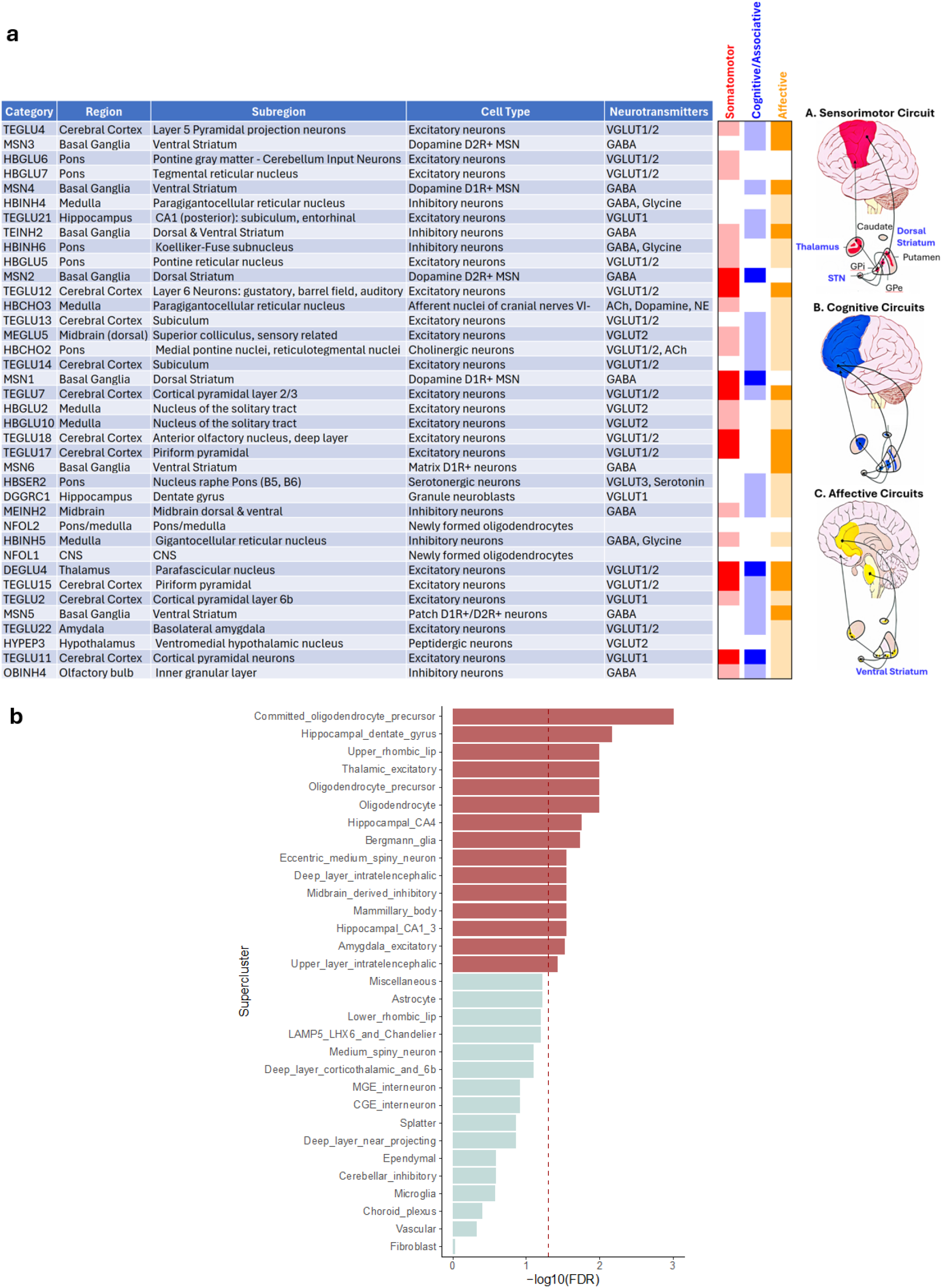
Tissue- and cell type-specific enrichment analyses of aggregated TD genetic risk using MAGMA. **A) Aggregated TD polygenic risk enrichment identifies mouse brain cell types** involved **in sensorimotor, cognitive/associative, and limbic cortico-striato-thalamo-cortical (CSTC) circuits**. Cell-type enrichment analysis was conducted using MAGMA, based on gene expression profiles from 265 cell types identified from single-cell RNA sequencing data of the mouse ^92^brain^93^. Thirty-eight cell types demonstrated significant enrichment of TD-associated genetic risk after FDR correction (FDR < 0.05). Each significantly enriched cell type is annotated with its anatomical location, structure, and associated neurotransmitter. Dark red, blue and orange bars indicate cell types involved in sensorimotor, cognitive/associative, and affective CSTC circuits, respectively. Light red, blue and orange bars indicate cell types involved the extended corresponding CSTC circuitry activities. **B) Enrichment of TD genetic risk across 31 human brain cell-type superclusters**. This analysis was based on single-nucleus RNA sequencing (snRNA-seq) data from the Brain Initiative Cell Census Network (BICCN), comprising over 3.3M nuclei from 100 distinct brain regions. Thirty-one superclusters were defined based on cell-type taxonomy, and their enrichment for TD genetic risk was assessed using MAGMA. Red bars indicate superclusters showing significant enrichment of TD genetic signals (FDR < 0.05), while light blue bars represent no significant enrichment. The dotted red vertical line marks the FDR threshold (FDR < 0.05). Abbreviations: MSNs, medium spiny neurons; GABA, Gamma-aminobutyric acid; VGLUT1 & VGLUT2, Vesicular glutamate transporter 1 & 2; Gly, Glycine; Ser, Serine; ACh, Acetylcholine; NE, Norepinephrine.

Analysis of human expression data in MAGMA provided further converging evidence. First, human bulk tissue RNA-seq data from GTEx^72^ evidenced significant enrichment in 7 of 11 brain tissues, including the basal ganglia (putamen, nucleus accumbens), limbic regions (amygdala, hippocampus), frontal and anterior cingulate cortex, and hypothalamus. No enrichment was observed in non-neuronal or peripheral nerve tissues (Supplementary Table 24, Supplementary Fig. 11a). We then analyzed 31 human brain cell-type superclusters derived from single-nucleus RNA-seq data from the Brain Initiative Cell Census Network^94^ (BICCN, >3.3M nuclei from 100 distinct brain regions) and found significant TD polygenic enrichment in 15 superclusters (Fig. 3b). Strikingly, both human and mouse data revealed a strong and significant enrichment of TD genetic signal in oligodendrocyte-lineage cells (precursors and mature oligodendrocytes), suggesting a role for myelination and glial support in TD pathophysiology. Furthermore, various disease-associated neuronal cell types were identified with enrichment of TD signal, such as the cerebral cortex excitatory neurons^95^ (deep−layer and upper-layer intratelencephalic), striatal eccentric MSNs^98^, hippocampal excitatory neurons^96^ (dentate gyrus, CA4, and CA1−3), thalamic excitatory neurons^97^, midbrain−derived inhibitory neurons^98^, and hypothalamus^99^ (mammillary body).

### Genetic correlation between TD and other traits

Using LDSC, we investigated genetic correlations (*r*_*g*_) between TD and 108 complex traits, including psychiatric, psychological, neurological, anthropomorphic, and other phenotypes of potential relevance to TDs (Fig. 4, Supplementary Table 25). After FDR correction, 46 traits showed significant correlations with TD. The strongest correlations were with other psychiatric disorders, including OCD (r_g_ = 0.51, P = 9.7 × 10^−49^), ADHD (r_g_ = 0.45, P = 2.6 × 10^−35^), ASD (r_g_ = 0.33, P = 4.0 × 10^−8^), anxiety (r_g_ = 0.32, P = 9.5 × 10^−12^), and MDD (r_g_ = 0.31, P = 3.7 × 10^−20^). Significant correlations were also observed for other potentially relevant traits, including suicide attempts (r_g_ = 0.33, P = 3.2 × 10^−10^) and neuroticism (r_g_ = 0.25, P = 1.9 × 10^−16^), cognitive traits including negatively correlated educational attainment (r_g_ = -0.16, P = 4.0 × 10^−10^), and substance use disorders and traits such as smoking (r_g_ = 0.17, P = 4.1 × 10^−9^) and cannabis use disorder (r_g_ = 0.25, P = 2.^5^ x 10^−5^). Although TD is considered to cross the boundaries between neurological and psychiatric disorders, of the neurological phenotypes studied, only migraine (with and without aura)^100^ was significantly associated with TD (r_g_ = 0.15, P = 2.3 × 10^−5^). Other significant associations of interest included asthma, sleep disorders such as insomnia and excessive daytime sleepiness, and measures of well-being, maltreatment, and disability.

**Fig. 4.**
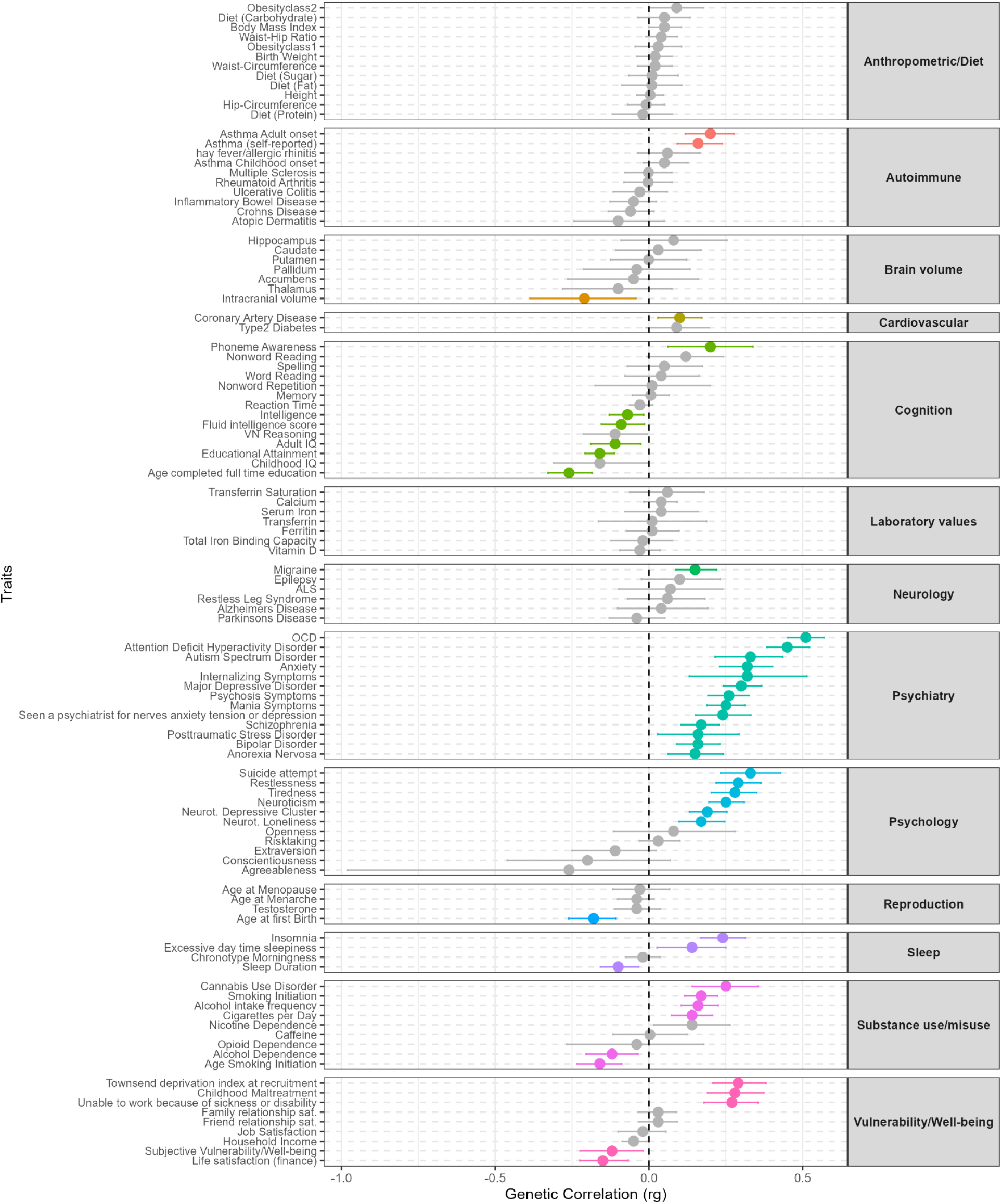
Genetic correlation between TD and 108 traits, including traits in anthropometry/diet, autoimmune, brain volume, cardiovascular, cognition, laboratory values, neurology, psychiatry, psychology, reproduction, sleep, substance use/misuse, and vulnerability/wellbeing. References and sample sizes of the corresponding GWAS studies can be found in Supplementary Table 25. The dots represent the genetic correlation (*r*_*g*_) estimated from LDSC, and the error bars represent the 95% confidence intervals. Colored points and bars indicate significant genetic correlations after FDR correction.

Since 3 out of 14 cohorts were originally ascertained for other psychiatric disorders, we conducted sensitivity analyses by excluding these 3 cohorts (TD_minusCOM subset) to address potential inflation of the genetic correlation estimates due to these cohorts (Supplementary Table 25, Supplementary Fig. 12). Genetic correlation estimates from the full TD GWAS and the TD_minusCOM subset GWAS were highly concordant (*Pearson’s r* = 0.95, *P* < 2.2 × 10^−16^), although linear regression revealed a slope greater than 1 (β = 1.16, *P* = 1.9 × 10^−5^), suggesting modest inflation of genetic correlation estimates when including these cohorts. This inflation was most pronounced for genetic correlations with psychiatric disorders. Despite this inflation attributable to the presence of co-occurring diagnoses, a substantial component of the observed pleiotropy reflects a shared underlying polygenic architecture.

## Discussion

In this largest TD GWAS to date, comprising 13,247 individuals with tic disorders ranging from TS to TTD, we identified six loci significantly associated with TD and substantially refined the understanding of the cellular and neuroanatomical basis of these disorders. Our results provide convergent genetic evidence implicating specific neuronal and glial cell populations within the CSTC circuits, prioritize novel risk genes for TD, replicate previously identified risk genes, and quantify the extensive genetic correlation between TD and other neuropsychiatric conditions.

A key finding of this GWAS is the association of TD genetic risk with specific cell types within the CSTC circuitry, a network long hypothesized to be central to TD pathophysiology^101^. Our analyses provide genetic support for this model and extend it by demonstrating that TD risk is not confined to a single functional domain. The enrichment of polygenic risk across somatomotor, cognitive/associative, and affective/limbic components of the CSTC network aligns with the clinical complexity of TD, which encompasses not only tics but also a high rate of co-occurring disorders, in particular, OCD and ADHD^10^. Crucially, within the striatum, our results point to both D1R+ and D2R+ MSNs as key substrates of TD genetic risk. The enrichment in D2R+ MSNs, the principal neurons of the indirect (movement-suppressing) pathway, aligns with the efficacy of the most common medications for tics, which are D2R antagonists such as aripiprazole, risperidone, and haloperidol. Conversely, the implication of D1R+ MSNs of the direct (movement-facilitating) pathway aligns with the recent development of novel therapeutic strategies, such as the D1 receptor antagonist ecopipam, which has shown promise in the treatment of tics^102^. Finally, additional polygenic enrichment was seen in layer 5 cortical pyramidal neurons and thalamic excitatory neurons of the parafascicular nucleus, a primary target for deep brain stimulation for refractory TS, genetically validating other key therapeutic and pathophysiological nodes in the circuit.

Notably, we also identified a strong enrichment in oligodendrocyte-lineage cells. This novel genetic finding is consistent with a growing body of evidence pointing to the importance of white matter integrity in TD. Diffusion imaging studies in both drug-naïve pediatric and adult TS patients have found microstructural alterations in white matter tracts connecting CSTC nodes^103–105^. Furthermore, mouse models of TS pathophysiology exhibit corresponding white matter abnormalities in these same circuits^106^. Our findings provide genetic support for these observations, suggesting that TD risk variants may act within oligodendrocytes to disrupt mylination^107^, thereby impairing the signaling efficiency and coordination of CSTC circuits. This result, along with previous work implicating astrocyte metabolism in TS^69^, points to a broader, integral role for glial cell dysfunction in the etiology of the disorder.

Integrative gene prioritization implicated in the TD GWAS highlighted genes central to neurodevelopment. The novel 13q21 locus implicates *PCDH9*, a protocadherin involved in synaptic adhesion and connectivity that has also been associated with autism and MDD^108^, while the 5p11-p12 signal implicates *HCN1*, a key regulator of neuronal excitability that has been linked to infantile epileptic encephalopathy^64,109^. In the frontal cortex, the HCN channel is regulated by the α2A noradrenergic receptor, which is the drug target of the α2 agonists guanfacine and clonidine, two first-line pharmacological treatments for TD and for ADHD, a commonly comorbid condition that shares genetic risk with TD. Notably, *HCN4*, another member of the HCN family, was previously linked to TS through elevated anti-HCN4 antibody levels in TS patients compared with controls and supported by a behavioral study in an animal model^91^. Heteromeric HCN1-HCN4 channels are crucial for generating rhythmic activity in heart and brain cells by regulating cell membrane excitability^92^.

Furthermore, the pleiotropic locus at 3p21 that is associated with TD harbors multiple candidate genes, including *NCKIPSD, WDR6, DALRD3, and CELSR3*. The convergence of evidence on this locus, associated in previous GWAS with OCD^55^, ADHD^110^, MDD^111^, and anorexia nervisa^112^ highlights it as a possible region of shared genetic risk that impacts circuit development across related disorders. Genes in this region play a role in synaptic signaling^51^, neuronal growth and migration^113^, and axonal targeting, potentially implicating these processes in the development of this complex neurodevelopmental disorder^114^.

The high clinical comorbidity in TD is mirrored by extensive genetic overlap with other complex traits, aligning with growing evidence for a shared genetic architecture across psychiatric disorders^115^. Our results quantify this overlap for TD, revealing strong positive genetic correlations with OCD, ADHD, suicide attempts, ASD, anxiety, and MDD. Together, these findings place TD within a transdiagnostic spectrum of neurodevelopmental and psychiatric conditions that share common biological mechanisms. Of note, genetic pleiotropy, while hypothesized, cannot be unequivocally determined from our data. Genetic correlations can result from mechanisms other than genetic pleiotropy, including mediation by one trait of the genetic liability of the other trait^116^.

Our data, combined with previous studies^55,117^, suggest that shared liability between TD and other psychiatric traits (in particular, OCD and ADHD) may converge on the CSTC circuits and cell types identified in our analyses, including D1/D2 MSNs and oligodendrocytes, which are integral to motor, cognitive, and affective processes. This interpretation is consistent with cross-disorder evidence implicating neurodevelopmental and synaptic pathways as core mediators of pleiotropic genetic effects^118^.

While this study’s scale is a major strength, we acknowledge its limitation to individuals of genetically inferred European-associated ancestry, which necessitates further investigation in diverse populations. The phenotypic heterogeneity across cohorts, though mitigated by high genetic correlations between ascertainment strategies, may have attenuated our power to detect additional loci. Furthermore, the reliance on adult transcriptomic reference data limits our ability to capture risk acting during prenatal or early postnatal windows. For example, *PCDH9* shows peak expression in the early postnatal brain before declining to lower levels in adulthood^40^, likely explaining why it was not prioritized by adult expression-based approaches.

In summary, the findings from this study advance the genetic understanding of TD in several important domains. Beyond locus discovery and risk gene prioritization, our analyses highlight specific cell types and brain circuits likely mediating genetic risk for TD. This cellular and anatomical specificity will likely provide a critical foundation for future mechanistic studies and for the development of targeted therapeutic strategies. Our PGS findings further demonstrate the predictive utility of TD genetic risk, which, while not yet clinically actionable, provides a framework for future risk stratification in the clinical setting.

## Data Availability

The meta-analyzed summary statistics will be made available from the Psychiatric Genomics Consortium download or upon reasonable request to the authors.

## Acknowledgements

We would like to thank all research participants and employees of all cohorts included in this study. This project was funded by the NIH grants R01 NS105746 to C.A.M., J.M.J., and P.P.; R01 NS102371 and Rosen Family Foundation to C.A.M. and J.M.J.; R01NS102371-01A1 and R01MH137220 to L.K.D.; the Evelyn F. and William L. McKnight Brain Institute to L.M.P. and P.G.R.; the NIH grant R01ES034554 to J.G.; National Health and Medical Research Council (NHMRC) EL1 Investigator Grant fellowship 2034743 to Z.F.G.; KNAW Royal Netherlands Academy of Science Professor Award (PAH/6635) to D.I.B.; NIH/NHGRI U01HG007269 to L.H.C.; the National Institute of Mental Health (R01MH137578) to M. Gandal and M.T.. The TIC-Genetics cohort was supported by grants from the National Institute of Mental Health (R01MH115958 to Gary A Heiman and Jay A Tischfield, R01MH115959 to Barbara J. Coffey, R01MH115960 to Alyssa Rosen, R01MH115961 to Samuel Kuperman, R01MH115962 to Donald Gilbert, R01MH115963 to Mathew State and A. Jeremy Willsey, and R01MH115993 to Samuel H. Zinner). Additional support for TIC Genetics was provided by Human Genetics Institute of New Jersey (to G.A.H. and J.A.T.), and the New Jersey Center for Tourette Syndrome and Associated Disorders (to G.A.H. and J.A.T.). We are also grateful to the NJCTS its long-standing suport of the TIC Genetics consortium. The BioVU cohort was funded by the NIH funded Shared Instrumentation Grant S10RR025141, the CTSA grants UL1TR002243, UL1TR000445, and UL1RR024975. The BioVU genomic data was supported by U01HG004798, R01NS032830, RC2GM092618, P50GM115305, U01HG006378, U19HL065962, R01HD074711. The iPSYCH team was supported by the Lundbeck Foundation (R102-A9118, R155-2014-1724, and R248-2017-2003), NIH/NIMH (1R01MH124851-01 to A.D.B.), and the Universities and University Hospitals of Aarhus and Copenhagen. The Danish National Biobank resource was supported by the Novo Nordisk Foundation. High-performance computer capacity for handling and statistical analysis of iPSYCH data on the GenomeDK HPC facility was provided by the Center for Genomics and Personalized Medicine and the Centre for Integrative Sequencing, iSEQ, Aarhus University, Denmark (grant to A.D.B.). ECC is supported by the Research Council of Norway (#274611) and South-Eastern Norway Regional Health Authority (#2021045), and is a member of the MRC Integrative Epidemiology Unit at the University of Bristol which is supported by the Medical Research Council and the University of Bristol (MC_UU_00032/1). Funding support for the Study of Addiction: Genetics and Environment (SAGE) was provided through the NIH Genes, Environment, and Health Initiative [GEI] (U01 HG004422); SAGE is one of the genomewide association studies funded as part of the Gene Environment Association Studies (GENEVA) under the NIH GEI. Assistance with phenotype harmonization and genotype cleaning, as well as with general study coordination, was provided by the GENEVA Coordinating Center (U01 HG004446). Assistance with data cleaning was provided by the National Center for Biotechnology Information. Support for collection of data sets and samples was provided by the Collaborative Study on the Genetics of Alcoholism (U10 AA008401), the Collaborative Genetic Study of Nicotine Dependence (P01 CA089392), and the Family Study of Cocaine Dependence (R01 DA013423). Funding support for genotyping, which was performed at the Johns Hopkins University Center for Inherited Disease Research, was provided by the NIH GEI (U01HG004438), the National Institute on Alcohol Abuse and Alcoholism,NIDA, and the NIHcontract “High Throughput Genotyping for Studying the Genetic Contributions to Human Disease” (HHSN268200782096C). The data sets used for the analyses described here were obtained from dbGaP (http://www.ncbi.nlm.nih.gov/projects/gap/cgibin/study.cgi?study_id=phs000092.v1.p1) through dbGaP accession number phs000092.v1.p.

## Author Contributions

C.A.M., J.M.S., D.Y., and P.P. designed the study. D.Y., N.I.S., Z.F.G., A.T., M.W.H., S. Shekhar, M. Tang, L.M.P., and F.I. conducted post-GWAS data analyses. D.Y., N.I.S., A.T., M.W.H., S. Shekhar, T.W.M., B.M., T.P., E.C.C., C.A., A.A., H.A., V.B., J. Ball, C.L.Barr, C. Barta, E.B., J.R.B., N.B., F.B., D.I.B., A.D.B., C.L.B., J.K.B., J. Buse, J.B., F.C., D.C., L.H.C., K.C., B.J.C., N.D., C.D., A.D., L.D., P.D., G.E., E.M.E., S.F., D.F., T.V.F., J.P.F., N.J.F., A.F., C.F., E.G., B.G., M.G., D.L.G., J. Glennon, D.G., M.A.G., E.L.G., J.G., D.F.G., A. Hartmann, A.H., T.H., B.H., L.D.H., I.H., P.J.H., H.J.H., A.Y.H., C. Huijser, D.I., P.J., J.J., C.J., S.J., A.S.K., M.K., C.K., J.K., I.K., N. Khalifa, Y.K., R.A.K., N.K., C.S.K., Y.K., A.K., S.K., J.F.L., B.L.L., H.L., C.L., M.L.L., M. Madruga-Garrido, I.A.M., O.M., W.M.M., S.M.M., M.D.M., D.M., P.M., A. Morer, N.M., K.M., A.M., L.M., T.M., P.N., B.M.N., E.L.N., K.S.O., M.S.O., S.S.P., D.L.P., K.P., G.P., C.P., D.P., P.J.W.P., J.P., R.R., M.R., V.R., A.R., G.R., S. Sandin, P.S., M.S., S. Schmitt, J.A.S., H.S.S., J.W.S., S.S., T.S., N.S., U.S., Z. Tarnok, O.T., M.T., J.A.T., Z.T., A.U., O.A.v.d.H., Y.D.v.d.W., M.C.V., D.J.V., A.V., G.B.W., B.W., T. Werge, J.W., T.W., Y.W., J.X., J.Y., C.Z., N.R.Z., S.H.Z., J.D.B., D.E.G., C.H., H.S., O.A., A.P., G.A.H., L.K.D., M.M., J.J.C., P.P., J.M.S., and C.A.M. provided samples and/or performed cohort-specific analyses. D.Y., N.I.S., Z.G., A.T., M.W.H., S. Shekhar, T.W.M., L.M.P., F.I., A.F., P.G.R., M.M., P.P., C.A.M., and J.M.S. wrote the paper and formed the core revision group. C.A.M., J.M.S., P.P., J.J.C., M.M., P.G.R., L.K.D., and M. Gandal supervised and directed this study. C.A.M., J.M.S., and P.P. obtained the funding for this study. All authors contributed to the discussion and approved the final version of the manuscript.

## Methods

### Ethics

All participants provided informed consent, and all studies obtained ethical approvals from local ethics review boards. Detailed ethical approval for each cohort is provided in Supplementary Note 2.

### Participating cohorts

The TD meta-analysis comprises 14 case-control cohorts of genetically inferred European-associated ancestry (Supplementary Table 1), consisting of 6 datasets from previous TD GWAS^1,19,36^ and 8 new cohorts, totaling 13,247 cases and 536,217 ancestry-matched controls. All cases were ascertained through semi-structured interviews, national health registries, or hospital biobanks. Seven cohorts were recruited from clinically ascertained cases diagnosed by semi-structured clinical interviews (N = 6,425; ICD-10 F95.2 or DSM-5 TS). One cohort included biobank participants identified by tic disorder algorithm^119^ (N = 319), and two cohorts (deCODE and MoBa) included national health registry-based cases with the F95.2 and F95.1 ICD-10 codes (N = 2,697). One additional national registry-based cohort (FinnGen) contributed individuals with any tic disorder (ICD-10 codes F95.2, F95.1, F95.0, F95.8, or F95.9; N = 293). The remaining three cohorts consisted of participants originally ascertained for studies focusing on another psychiatric disorder who had a comorbid diagnosis of tic disorder (ICD-10 F95.2, F95.1, F95.0, F95.8, or F95.9; N = 3,513). Ancestry-matched controls were recruited as either unscreened individuals or individuals without known tic disorders. Detailed descriptions of each cohort’s ascertainment, study design, genotyping, quality control, imputation, and GWAS procedures are reported in Supplementary Note 2.

### Meta-analysis

We conducted an inverse variance-weighted meta-analysis across all 14 cohorts (13,247 cases and 536,217 controls) using a fixed-effects model implemented in METAL v.2011-03-25^120^. Summary statistics from individual cohort GWAS were harmonized to the hg19 reference genome, and strand orientation was aligned to the HRC panel. Variants with an effective sample size > 10,000 and ≥ 5,000 cases were retained, yielding 7,080,961 SNPs for analysis.

We quantified between-cohort heterogeneity using Cochran’s I^2^ statistic and assessed test statistic inflation (genomic control factor λ and λ_1000_) for each cohort’s GWAS and the meta-analyses. LD Score Regression (LDSC v1.0.1)^121^ was used to partition inflation into components due to polygenicity versus confounding, using high-quality common variants (INFO > 0.9, MAF > 0.01).

To evaluate heterogeneity by study design, we conducted stratified meta-analyses according to ascertainment method and phenotypic inclusion criteria (Supplementary Tables 2-3). Two meta-analyses were conducted on the cohorts stratified by ascertainment methods: 1) cohorts ascertained with semi-structured interviews or validated questionnaire (SQ), including TAAICG_610K, TAAICG_Omni, TAAICG_fam, TAAICG_GSA, TAAICG_PsychChip, TIC_Genetics, and EMTICS, with a total of 6,425 cases and 12,580 controls; and 2) cohorts with cases recruited from national registries (NR), namely MoBa, deCODE, FinnGen, NORDiC_SWE, EGOS, and iPSYCH, with 6,503 cases and 522,052 controls. To investigate association heterogeneity across phenotypic subgroups, three additional meta-analyses were performed on cohorts stratified by the phenotypic inclusion criteria: 1) TS subset contains cohorts with at least 95% of cases with TS (meeting F95.2 ICD 10 code or Tourette Syndrome in DSM-5 or DSM-IV-TR), which comprised TAAICG_610K, TAAICG_Omni, TAAICG_fam, TAAICG_GSA, TAAICG_PsychChip, TIC_Genetics, and TS_EURO, totaling 6,425 cases and 12,580 controls. 2) TD subset contains cohorts with both TS and PMVTD cases meeting F95.2 or F95.1 (deCODE) or cases with any tic disorder (BioVU, Moba, and FinnGen), resulting in 3,309 cases and 492,279 controls. 3) TD_comorbid subset contains cohorts with any TD cases that were primarily ascertained for a study on another comorbid psychiatric disorder, encompassing iPSYCH, NORDiC_SWE, and EGOS, with 3,513 cases and 31,358 controls. Genetic correlations among stratified meta-analyses were estimated using LDSC.

#### Genomic structural equation modeling (Genomic SEM)

To model the shared genetic architecture across the three stratified subsets, we performed confirmatory factor analysis (CFA) using Genomic SEM^122^. A one-factor model (without SNP effects) was specified under unit variance identification, fixing the latent factor variance to 1 and freely estimating the trait loadings. Model parameters were estimated using the Diagonally Weighted Least Squares (DWLS) method. Detailed instruction can be found in the “Models without individual SNP effects” tutorial on the GenomicSEM GitHub website (https://github.com/GenomicSEM/GenomicSEM/wiki/3.-Genome%E2%80%90wide-Models).

#### Estimation of the number of TD causal variants using MiXeR

We used MiXeR v1.3^123^ to characterize the polygenic architecture of TD. MiXeR models genome-wide association summary statistics using a univariate Gaussian mixture model, in which common variants are assumed to comprise a mixture of causal and noncausal effects. Polygenicity was quantified as the number of causal variants that account for 90% of the SNP-based heritability of TD.

#### Conditional and joint analysis (GCTA-COJO)

We used the GCTA-COJO method^124^ to identify independent genetic signals within significant TD loci. This approach performs conditional and joint multiple-SNP analysis to detect additional (i.e., secondary) association signals that may not be identified through single-SNP analyses. Independent SNPs were identified using the default stepwise model selection procedure. Linkage disequilibrium (LD) reference data were derived from 73,005 individuals in the QIMR Berghofer Medical Research Institute genetic epidemiology cohort. A distance of 10 Mb was assumed for complete LD, and a genome-wide significance threshold of *P* < 5.0 × 10^−8^ was applied.

### Fine mapping

To identify potential causal variants among the six genome-wide significant (GWS) loci, we performed Bayesian fine-mapping using PAINTOR^66^. This method integrates GWAS association strength, LD structure, and functional annotations to estimate posterior probability of causality of each SNP within the linkage disequilibrium (LD) block surrounding the GWS loci. We considered SNPs with GWAS *P* < 1.0 × 10^−6^ and *r*^2^ > 0.5 within ±1 Mb of SNPs with GWAS *P* < 5.0 × 10^−8^. Functional annotations (Supplementary Table 10) included gene elements, brain-specific enhancers, brain DNase I hypersensitive sites (DHS), transcription factor binding sites (TFBS), promoters, and constraint regions in mammals and primates. We assumed no more than 2 causal variants per locus and considered any SNP with a posterior probability > 0.5 as a potential causal variant.

### SNP-heritability and genetic correlation estimation

SNP-based heritability was estimated using LDSC^121^ with 1000 Genomes European LD scores^125^. Liability-scale heritability was computed assuming a 0.8% population prevalence for TS and 1.5% for TD and TD_comorbid subsets. The effective sample-size weighting across cohorts was used, and the corresponding sample prevalence was fixed at 50%.

Genetic correlations (r_g_) were estimated using bivariate linkage-disequilibrium score regression. To ensure robust estimation, we excluded GWAS with fewer than 5,000 cases (for case-control studies), total sample sizes below 10,000 (for quantitative traits), or fewer than 500,000 SNPs overlapping with the TD GWAS. A total of 108 published GWAS were analyzed to assess for genetic correlations with TD. These traits were grouped into 13 categories: anthropometry/diet, autoimmune, brain volume, cardiovascular, cognition, laboratory values, neurology, psychiatry, psychology, reproduction, sleep, substance use/misuse, and vulnerability/well-being. Summary statistics were obtained from LD hub ^126^, the Psychiatric Genomics Consortium (PGC) (https://pgc.unc.edu/for-researchers/download-results/), or through direct request to study authors. Detailed information is provided in Supplementary Table 25. To account for multiple testing, we applied the Benjamini-Hochberg correction^127^ to calculate the false-discovery rate (FDR).

To evaluate the potential impact of comorbid cases from three cohorts originally ascertained for other psychiatric disorders, we performed a sensitivity analysis using a subset of TD samples excluding these cohorts (TD_minusCOM subset). We re-estimated genetic correlations between TD_minusCOM and the 108 traits (*r*_*g*__minusCOM) and assessed the concordance between *r*_*g*_ from the full meta-analysis and *r*_*g*__minusCOM.

### Polygenic scores

For the polygenic score (PGS) analyses, we calculated individual PGS in four cohorts (EMTICS, TAAICG_610, TAAICG_Omni, and TIC_Genetics) for which individual-level genotype data were available and which contained >500 cases and >500 controls. For the discovery sample, we first conducted a leave-one-out meta-analysis without the target cohort and kept the SNPs with INFO > 0.8 and MAF > 0.01 in both the leave-one-out discovery sample and the target cohort. Secondly, we estimated posterior SNP effect sizes in PRScs^128^ under continuous shrinkage (CS) priors using the leave-one-out summary statistics and an LD matrix calculated from the HRC European reference panel. The estimated posterior SNP effect sizes were then used to calculate a weighted sum of allele dosages as the PGS in the target sample. The PGS were then standardized within each target cohort for cross-cohort comparison by adjusting for PCAs and number of SNPs used for PGS. We evaluated the predictive power using Nagelkerke’s R2 and plots of odds ratios over score deciles. Both R2 and odds ratios were estimated in logistic regression analyses, adjusting for ancestry principal components.

### Gene-based Analyses and Gene Prioritization

To prioritize the genes most likely contributing to TD risk, we utilized a comprehensive approach combining positional and functional gene-based analyses. This involved integrating functional genomic annotations, transcriptomic analysis, Mendelian Randomization (MR) analysis, and colocalization analysis.

#### Positional Gene Mapping

Positional gene mapping was conducted using two distinct approaches: multivariate Set-Based Association Test (mBAT-combo)^70^ and Multi-marker Analysis of GenoMic Annotation (MAGMA)^129^.

The mBAT-combo method integrates mBAT and fastBAT test statistics via a Cauchy combination method, enabling the incorporation of diverse test statistics without prior knowledge of their correlation structure. This maximizes statistical power regardless of masking effects at specific loci. We implemented mBAT-combo within GCTA version 1.94.1^124^. The European subsample (N = 503) from Phase 3 of the 1000 Genomes Project was used as the linkage disequilibrium (LD) reference panel^125^, with a fastBAT default LD cut-off of 0.9. Our analysis focused on a gene list of 19,899 protein-coding genes mapped to the genome build hg19.

MAGMA incorporates LD structure and utilizes a multiple regression approach to detect the multi-marker effects^129^. We employed MAGMA v1.10 to identify annotated genes carrying significant TD risk variation from the summary statistics. Gene annotations for 19,175 gene units were obtained from a previously published source (https://github.com/jbryois/scRNA_disease/raw/refs/heads/master/Code_Paper/Data/NCBI/NCBI37.3.gene.loc.extendedMHCexcluded) and corresponded to protein-coding genes from Ensembl version 87. Each gene unit was defined by an interval 35 kilobases upstream of the transcription start site to 10 kilobases downstream of the transcription stop site.

#### Functional Gene Mapping

Five distinct approaches were employed for functional gene mapping: E-MAGMA, TWAS_JTI, TWAS+COLOC, SMR+HEIDI, and H-MAGMA.

**E-MAGMA** is a modified MAGMA approach that assigns tissue-specific cis-eQTLs (FDR<0.05) to their putative genes for multi-marker regression analysis, rather than relying solely on positional SNP assignment^130^. In this study, we assigned SNPs defined as eQTLs in GTEx v8 in at least one brain tissue from GTEx v8.

**Transcriptome-Wide Association Study (TWAS) using Joint Tissue Imputation (JTI) (TWAS_JTI)** was conducted using summary statistics from the TD GWAS. Precomputed JTI weights from 49 GTEx v8 tissues were utilized to impute genetically regulated gene expression and perform an association test^131^. JTI enhances imputation accuracy, particularly for tissues with limited availability, such as the brain, by jointly modeling across tissues and leveraging shared genetic architecture. Gene-level associations from each tissue were combined using the Aggregated Cauchy Association Test (ACAT) ^132^, which transforms individual p-values into Cauchy-distributed variables and aggregates them via weighted summation.

**TWAS with Colocalization Analysis (TWAS+COLOC)** integrated GWAS summary statistics with gene expression prediction models to identify associations between gene expression and TD risk^73^. We used brain gene expression weights from the PsychENCODE project^133^ and LD information from the 1000 Genomes Project Phase 3^125^. These reference panels facilitate the estimation of the genetic component of gene expression, which is then tested for association with TD using GWAS summary data. For colocalization analyses, we employed the COLOC R package^74,75^, implemented using TWAS FUSION. This Bayesian method calculates posterior probabilities (PP) to assess whether individual lead SNPs within a significant TWAS locus represent an independent signal (PP3) or shared signals (PP4) affecting both transcription and TD. This integrative approach enhances the detection of gene-trait associations by combining genetic association data with gene expression prediction models, providing insights into the genetic architecture of TD.

**Summary data-based Mendelian Randomization (SMR) with HEIDI (SMR+HEIDI)** was conducted to identify genes whose expression levels are associated with TD through pleiotropy^76^. We utilized eQTL meta-analysis summary statistics from European populations, specifically whole blood data from eQTLGen^77^ and data from five nervous system tissues (basal ganglia, cerebellum, cortex, hippocampus, and spinal cord) from MetaBrain^134^. To distinguish between linkage and pleiotropy, the Heterogeneity in Dependent Instruments (HEIDI) test was performed alongside SMR, evaluating effect size heterogeneity between the GWAS and eQTL summary statistics. A HEIDI P-value of 0.05 was used to determine the likelihood of linkage influencing the observed association. This combined approach aids in identifying causal genes associated with TD by distinguishing pleiotropic effects from linkage.

H-MAGMA was a modified MAGMA approach that incorporates fetal brain Hi-C data (paracentral cortex from gestation week 17∼18, N=3), adult brain Hi-C data (Dorsolateral Prefrontal Cortex (DLPFC) from individuals aged 36, 44 and 64 years, N=3), and neuronal and astrocytic Hi-C data derived from human iPSCs (N=2)^135^.

A gene demonstrating significance in at least one positional test (the left panel) and at least 3 functional tests is classified as a high-confidence TD risk gene.

### Tissue and cell type specificity test

To investigate the tissue and cell-type enrichment of genetic associations with tic disorder (TD), we performed tissue and cell-type enrichment analyses using four datasets. First, we followed the protocol established by Bryois et al.^136^, obtaining the gene sets marking specific tissues and cell types from this publication (https://github.com/jbryois/scRNA_disease). These datasets included: 1) 37 GTEx testable tissues^137^; 2) 39 broad mouse neuronal populations; and 3) 265 specific mouse neuronal populations. The last two datasets were both derived from single-cell RNA sequencing of whole mouse brains^138^.

The fourth dataset is the human single-nucleus RNA-seq (snRNA-seq) data from the Brain Initiative Cell Census Network (BICCN) (available at CELLxGENE: https://cellxgene.cziscience.com/collections/283d65eb-dd53-496d-adb7-7570c7caa443), comprising 3,369,219 nuclei from 106 dissections across forebrain, midbrain, and hindbrain, spanning prenatal, early postnatal, and adult stages. This dataset spans prenatal, early postnatal, and adult stages. We used 31 superclusters defined by Siletti et al.^94^ for enrichment analysis. Pseudo-bulk differential expression analysis was conducted by aggregating gene counts per donor (excluding donor H18.30.001 due to low cell count) using DESeq2^139^ (likelihood ratio test). Genes with FDR-adjusted *P* < 1.0 × 10^-5^ were retained as cell-type-specific markers. A specificity index was calculated as the mean expression of a gene in a target cell type divided by its mean expression across all cell types, using a modified generate_celltype_data.r function from EWCE v1.10.2^140^.

For all datasets, enrichment was assessed using MAGMA^141^. SNPs were mapped to genes within a 10-kb upstream and 1.5-kb downstream window, with linkage disequilibrium adjusted using the 1000 Genomes Phase 3 European panel^125^. Cell types with FDR-adjusted *P* < 0.05 were deemed significantly associated with TD. For GTEx and broad mouse neuronal cell types, we also applied LDSC^121^, requiring FDR-adjusted *P* < 0.05 from both MAGMA and LDSC for significance.

### Review of pleiotropy and prior TD genetic findings

To assess the pleiotropic effects of potential TD risk genes identified in this manuscript, we searched the NHGRI-EBI GWAS Catalog^142^ (https://www.ebi.ac.uk/gwas/) and PubMed (https://pubmed.ncbi.nlm.nih.gov/). We queried for associations between these genes and other psychiatric, psychological, or neurodevelopmental disorders.

To identify genes previously reported in association with TD, we searched PubMed using the keywords “Tourette” or “tic disorder” and “gene”. We included findings from genome-wide studies of both common and rare variants but excluded candidate gene studies.

To validate these previously reported findings in our current study (Supplementary Tables 8-9), we used two approaches. For genes identified from rare-variant studies, we extracted the lowest P-value from our meta-analysis for any SNP within the gene’s transcription region (plus 50kb flanking regions on both ends). For previously reported GWS loci, we reported the P-value of the specific previously reported lead SNP in our data.

### Chromatin Interaction Analysis

To characterize chromatin interactions linking TD-associated risk variants to target genes, we examined chromatin conformation profiles across five brain-relevant biological contexts. These included fetal and adult cortical Hi-C datasets generated by Giusti-Rodríguez et al.^143^, as well as three induced pluripotent stem cell (iPSC)–derived neural cell types: neural progenitor cells (NPCs), excitatory glutamatergic neurons (EGNs), and medium spiny neurons (MSNs). Details of fetal and adult cortical Hi-C data generation and processing are described in Giusti-Rodríguez et al. (2019). Briefly, chromatin contact matrices were constructed at 1 Mb, 100 kb and 40 kb resolution from fetal (n = 3) and adult (n = 3) cortex samples, aligned to the hg19 reference genome. For iPSC-derived cell types, Hi-C experiments were performed in duplicate for each of NPCs, EGNs, and MSNs, and chromatin interaction analyses were conducted separately for each differentiated cell type. Genomic coordinates for iPSC-derived datasets were initially mapped to hg38 and subsequently converted to hg19 using the UCSC LiftOver tool, with a minimum mapping ratio of 0.70, minimum query hit size of 50 bp, minimum target chain size of 0, and allowing multiple output regions.

Frequently interacting regions (FIREs) were identified following the framework described by Schmitt et al.^144^ and implemented by Giusti-Rodríguez et al. (2019) for fetal and adult cortex samples. For iPSC-derived NPCs, EGNs, and MSNs, FIREs were detected using FIREcaller^145^ with identical parameter settings. FIRE detection was based on 40 kb resolution Hi-C contact matrices for each chromosome. For each 40 kb genomic bin, the total number of cis intra-chromosomal interactions within a genomic distance of 15–200 kb was calculated. Systematic biases related to effective fragment length, GC content, and mappability were corrected using HiCNormCis. Bins overlapping the major histocompatibility complex (MHC) region were excluded, resulting in 64,222 bins retained for analysis. The normalized total number of cis interactions was defined as the FIRE score. FIRE scores were quantile -normalized across samples using the R package preprocessCore to reduce batch effects, log2-transformed (log2[FIRE score + 1]), and standardized to Z-scores (mean = 0, standard deviation = 1). Bins with one-sided *P* < 0.05 were designated as FIREs.

Long-range chromatin interactions in fetal and adult cortex were identified using a combined Fit-Hi-C^146^ and FastHiC^147^ framework, as described by Giusti-Rodríguez et al. (2019). For iPSC-derived NPCs, EGNs, and MSNs, chromatin loops were detected using HiCCUPS^148^. Analyses were performed on 10 kb resolution contact matrices, evaluating genomic bin pairs separated by 30 kb to 2 Mb. Interactions overlapping centromeric regions, ENCODE blacklist regions, the MHC locus, or low-quality bins were excluded. High-confidence regulatory chromatin interactions (HCRCI) were defined using a Bonferroni-adjusted significance threshold of *P* < 1.16 × 10^−10^, corresponding to α = 0.005 across 42,985,244 tested 10 kb bin pairs. This global background modeling framework preferentially enriches for long-range enhancer–promoter interactions. Regulatory interactions were further annotated by intersecting loop anchors with promoter and enhancer elements using data from ENCODE Phase III regulatory elements (cCREs)^149^.

Independent genomic loci were mapped to HCRCI anchors if they were located within ±5 kb of either interaction anchor. Genes were assigned to regulatory interactions by extending gene bodies by ±5 kb and intersecting these extended regions with loop anchors using a custom Python-based pipeline. Genes meeting these criteria were considered linked to regulatory chromatin interactions. Protein-coding genes expressed in the human brain were defined using GENCODE v45 annotations^150^. The ±5 kb flanking window was selected to account for the resolution of loop anchors and to capture proximal regulatory contacts. All analyses were conducted separately for each cell type and developmental stage using R version 4.2 and Python.

