## Supplementary Notes and Figures for "Genome-Wide Association Analysis of Tic Disorders Reveals 6 Independent Risk Loci and Highlights Tic-Associated Cell Types and Brain Circuitry"

**Supplementary Notes**


| | | |
|---|---|---|
| Supplementary Note 1 | Cohort/Consortium-specific author lists | 2 |
| Supplementary Note 2 | Ethics and sample descriptions | 4 |
| Supplementary Note 3 | Gene-based tests | 17 |
| Supplementary Note 4 | Tissue/cell-type enrichment analyses | 18 |


**Supplementary Figures**


| | | |
|---|---|---|
| Supplementary Figure 1 | Quantile-Quantile (Q-Q) plots | 20 |
| Supplementary Figure 2 | Regional association plot and forest plot for SNP rs113907874 | 21 |
| Supplementary Figure 3 | Regional association plot and forest plot for SNP rs78995317 | 22 |
| Supplementary Figure 4 | Regional association plot and forest plot for SNP rs6768597 | 23 |
| Supplementary Figure 5 | Regional association plot and forest plot for SNP rs11720964 | 24 |
| Supplementary Figure 6 | Functional genomic architecture of the tic disorder (TD) genome-wide significant locus at chromosome 3p21 | 25 |
| Supplementary Figure 7 | Regional association plot and forest plot for SNP rs167184 | 26 |
| Supplementary Figure 8 | Regional association plot and forest plot for SNP rs8185387 | 27 |
| Supplementary Figure 9 | Decile plot of odds ratios by TD PRS within each decile for four cohorts and their meta-results | 28 |
| Supplementary Figure 10 | Enrichment of TD GWAS signals across 35 cell-type clusters from the mouse brain | 29 |
| Supplementary Figure 11 | Tissue and cell-type enrichment of TD genetic signals | 30 |
| Supplementary Figure 12 | Concordance of genetic correlations with and without cohorts ascertained for other psychiatric disorders | 31 |

**References** 32

**Supplementary Note 1: Cohort/Consortium-specific author lists**

TAAICG

Dongmei Yu, Zachary Gerring, Tyne W. Miller-Fleming, Luz M. Porras, Franjo Ivankovic, Cathy L. Barr, James R. Batterson, Fortu Benarroch, Cathy L. Budman, Danielle Cath, Laura Domènech, Emily Gantz, Marco A. Grados, Erica L. Greenberg, Luis Diego Herrera-Amighetti, Alden Y. Huang, David Isaacs, Joseph Jankovic, James F. Leckman, Christine Lochner, Irene A. Malaty, William M. McMahon, Benjamin M. Neale, Erika Nurmi, Michael S. Okun, David L. Pauls, Danielle Posthuma, Mary Robertson, Guy Rouleau, Paul Sandor, Joshua A. Senior, Harvey S. Singer, Lea K. Davis, Paola Giusti-Rodríguez, Peristera Paschou, Jeremiah M Scharf, Carol A Mathews

TS_EURO

**EMTICS:** Apostolia Topaloudi, Sudhanshu Shekhar, Alan Apter, Valentina Baglioni, Juliane Ball, Noa Benaroya-Milshtein, Judith Buse, Francesco Cardona, Andrea Dietrich, Blanca Garcia-Delgar, Marianthi Georgitsi, Tammy Hedderly, Isobel Heyman, Pieter J. Hoekstra, Chaim Huijser, Marcos Madruga-Garrido, Marieke D Messchendorp, Pablo Mir, Astrid Morer, Norbert Müller, Kirsten Müller-Vahl, Alexander Münchau, Peter Nagy, Kerstin Plessen, Cesare Porcelli, Renata Rizzo, Veit Roessner, Tamar Steinberg, Zsanett Tarnok, Marta Correa Vela, Jin Yin, Peristera Paschou

**TS-EUROTRAIN/TSGeneSEE:** Christos Androutsos, Csaba Barta, Entela Basha, Dorret I. Boomsma, Jan K. Buitelaar, Christel Depienne, Andrea Dietrich, Petros Drineas, Gudmundur Einarsson, Siyan Fan, Jakub P Fichna, Natalie J. Forde, Abel Fothi, Marianthi Georgitsi, Jeffrey Glennon, Daniel F. Gudbjartsson, Andreas Hartmann, Bastian Hengerer, Pieter J. Hoekstra, Piotr Janik, Cathrine Jespersgaard, Ahmad Seif Kanaan, Mira Kapisyzi, Iordanis Karagiannidis, Anastasia Koumoula, Shanmukha S Padmanabhuni, Geert Poelmans, Petra J. W. Pouwels, Joanna Puchala, Natalia Szejko, Urszula Szymanska, Olafur Thorarensen, Zeynep Tümer, Odile A. van den Heuvel, Ysbrand D. van der

Werf, Dick J. Veltman, G. Bragi Walters, Joanna Widomska, Tomasz Wolanczyk, Yulia Worbe, Cezary Zekanowski, Nuno R. Zilhäo, Hreinn Stefansson, Peristera Paschou

TIC Genetics

Alan Apter, Juliane Ball, Noa Benaroya-Milshtein, Danielle Cath, Keun-Ah Cheon, Barbara J. Coffey, Andrea Dietrich, Erik M Elster, Dana Feldman, Thomas V. Fernandez, Carolin Fremer, Donald L. Gilbert, Danea Glover, Tammy Hedderly, Isobel Heyman, Pieter J. Hoekstra, Hyun Ju Hong, Chaim Huijser, Christina Kappler-Friedrichs, Young Shin Kim, Robert A. King, Nadine Kirchen, Carolin Sophie Klages, Yun-Joo Koh, Samuel Kuperman, Bennett L. Leventhal, Holan Liang, Maria Loreta Lopez, Osman Malik, Marieke D Messchendorp, Dararat Mingbunjerdsuk, Pablo Mir, Astrid Morer, Kirsten Müller-Vahl, Alexander Münchau, Laura Muñoz-Delgado, Tara Murphy, Kerstin Plessen, Veit Roessner, Alyssa Rosen, Simon Schmitt, Sara Sopena, Zsanett Tarnok, Meitar Timmor, Jay A. Tischfield, Anne Uhlmann, Ana Vigil-Pérez, Belinda Wang, Jinchuan Xing, Samuel H. Zinner, Gary A. Heiman

BioVU

Tyne W. Miller-Fleming, Emily Gantz, David Isaacs, Lea K. Davis

deCODE

Gudmundur Einarsson, Daniel F. Gudbjartsson, Olafur Thorarensen, G. Bragi Walters, Hreinn Stefansson

MoBa

Elizabeth C. Corfield, Helga Ask, Alexandra Havdahl, Martin Scheiene, Ole Andreassen

FinnGen

Teemu Palviainen, Jaakko Kaprio, Aarno Palotie

iPSYCH

Nora I. Strom, Anders D. Børglum, Jonas Bybjerg-Grauholm, Jakob Grove, Sandra Melanie Meier, Thomas Werge, Manuel Mattheisen

NORDiC_SWE

Matthew W. Halvorsen, Kevin Sean O'Connell, James J. Crowley

EGOS

Behrang Mahjani, Niklas Dahl, Seulgi Jung, Najah Khalifa, Sven Sandin, Joseph D. Buxbaum, Dorothy E. Grice, Christina Hultman

PGC-CDG

Jordan W. Smoller

**Supplementary Note 2: Ethics and Sample Descriptions**

In the subsequent section, we describe each study included in the tic disorder (TD) meta-analysis. The studies are listed in alphabetical order. The header of each cohort includes the study identifier, country or site name, and relevant PubMed ID(s) if there have been previous publication(s) in connection with the data. The samples in all studies are of European ancestry only. Supplementary Table 1 provides an overview of the sample sizes, the numbers of included SNPs, the reference panel, and the genome-wide association analysis study (GWAS) analysis method/tool utilized, and the $\lambda_{1000}$ estimate for each study. All participants aged 18 and older gave informed consent. The individuals under 18 gave assent after a parent signed a consent form on their behalf. The research project was approved by the Ethics Committees of each participating site.

BioVU | USA | 18500243

BioVU is a large biobank at Vanderbilt University Medical Center in Nashville, TN. Participants who opt in to the BioVU program permit their leftover blood samples from routine clinical visits to be banked and genotyped for research purposes. These samples are linked to de-identified electronic health records for more than 300,000 individuals. Genotyping data for 94,474 BioVU individuals was performed using the Illumina MEGAEX array as previously described[1]. TD cases were selected by identifying BioVU individuals with a tic disorder ICD9 code or the presence of TD keywords within the medical record as previously described. In summary, TD cases identified by ICD9 code required two instances of a tic disorder code (307.2 Tics, 307.20 Tic Disorder NOS, 307.21 Transient tic disorder, 307.22 Chronic motor or vocal tic disorder, 307.23 Tourette's disorder, 333.3 Tics of organic origin). TD cases were excluded if they received an ICD9 code for motor disorders (333.6 Genetic torsion dystonia, 333.7 Acquired torsion dystonia, 333.72 Acute dystonia due to drugs, 333.79 Other acquired torsion dystonia, 333.8 Fragments of torsion dystonia, 333.82 Orofacial dyskinesia, 333.85 Subacute dyskinesia due to drugs, 333.89 Other fragments of torsion dystonia). Alternatively, TD cases could also be identified by the single presence of the following keywords within the medical records: "motor tic", "vocal tic", "Tourette", or "tic disorder". Each TD case was chart-reviewed by a clinician to confirm the TD diagnosis. Controls were age and sex-matched to TD cases and were required to have no instances of TD ICD9 codes, motor disorder ICD9 codes, or TD keywords within their medical records. Controls were additionally matched to the TD cases based on ancestry using Principal Component Analysis (PCA)[3]. SAIGE was used to perform a GWAS on 319 TD cases and 1,585 matched controls using covariates for sex, age, and PC1-10[4].

deCODE | Iceland | 28319091

The summary statistics for the tic disorder GWAS on the individuals from Iceland were provided by deCODE genetics[5]. This dataset encompasses 1,998 cases (1,097 cases with TS and 901 cases with chronic tics) and 134,508 unscreened population-matched controls. All cases met the criteria of ICD-10 code F95.2 or F95.1, or DSM-5 criteria for Tourette syndrome or Chronic Tics derived from a self-report questionnaire. Population samples without attention-deficit/hyperactivity disorder (ADHD), autism spectrum disorder (ASD), obsessive-compulsive disorder (OCD), or TS/TD were used as matching controls. Both cases and controls were genotyped at deCODE genetics using Illumina SNP arrays, with subsequent quality control and imputation using a previously described method[6]. The GWAS was conducted with a logistic regression model incorporating sex, county of origin, age, and the genetic relationship matrix as covariates. SNPs with INFO score <0.8 or minor allele frequency <0.01 were excluded for meta-analysis.

EGOS | Sweden | 31907560

We gathered genotype data for 341 cases and 4,024 controls in Sweden. The samples were genotyped using the Infinium Global Screening Array (GSA) or the Omni Express Exome array. Specifically, cases were sourced from three distinct studies: the EGOS cohort (Epidemiology and Genetics of Obsessive-Compulsive Disorder and Chronic Tic Disorders in Sweden; n=241; GSA)[7], the PAGES cohort (Population-Based Autism Genetics & Environment Study; n=53; OmniExpressExome)[8], and the UTC (Uppsala Tourette Cohort; n=47; OmniExpressExome)[9]. Control data were sourced from the LifeGene cohort (n=1,444; GSA)[10] and the Gaugler et al. study (n=2,580; OmniExpressExome)[11].

For the Gaugler et al. controls, we utilized data that had already undergone quality control (QC)[11]. For the remaining cohorts, our QC steps included removing individuals with a non-call rate greater than 0.05, conducting sex discrepancy tests, and assessing heterozygosity, particularly excluding individuals more than three standard deviations from the mean. Additionally, SNPs with a non-call rate greater than 0.05 were removed. For individuals of European ancestry, we applied a minor allele frequency (MAF) threshold greater than 0.01 and a Hardy-Weinberg Equilibrium threshold of $1.0 \times 10^{-10}$.

Using the McCarthy tool, we matched the SNPs to the HRC database. Post-QC, we merged the datasets of cases and controls genotyped on OmniExpressExome and those on GSA separately. Imputation of this data was performed using the Michigan Imputation Server with specific settings: the reference panel HRC r1.1 2016 (GRC37/hg19), phasing with Eagle v2.4, and a mixed population model.

After imputation, we further filtered SNPs, applying an imputation quality score ($r^2$) threshold of 0.81 and maintaining the MAF threshold at 0.01. The datasets were then merged based on common SNPs, followed by additional QC steps. These included removing duplicate SNPs, excluding closely related individuals (pihat > 0.2), and eliminating SNPs with a missingness rate in any cohort greater than 0.02. We also removed SNPs with a minor allele frequency of zero in at least one cohort and those with a (max – min) allele frequency difference greater than 0.2 across all individuals (cases and controls) and greater than 0.1 across merged cases and merged controls. Furthermore, SNPs with a (max – min) allele frequency difference greater than 0.03 in the controls and those with a fixation index ($F_{st}$) larger than 0.005 between control groups were also removed. Finally, we conducted a genome-wide association study using LifeGene controls as cases and Gaugler et al. controls as controls, removing variants with a p-value < 1e-4. This step, specifically designed for control groups, was crucial for identifying and excluding any spurious associations that could arise due to latent biases or technical artifacts.

The resulting data set, after the aforementioned quality control steps, included 311 cases and 3,952 controls, encompassing a total of 5,643,843 SNPs. Subsequently, we employed GemTools to exclude individuals with non-European ancestry. This refinement led to a final cohort comprising 284 cases and 3,703 controls, with a reduced SNP count of 5,545,509. For each case, we matched four controls based on the first two principal components (PCAs), sex, and the genotyping platform used. This resulted in a balanced dataset of 284 cases and 1,136 controls.

Finally, we conducted a genome-wide association study using logistic regression analysis in PLINK, incorporating the first six PCAs as covariates to account for ancestry and control for population stratification. These PCAs were derived using PLINK after conducting linkage disequilibrium (LD) pruning of the SNPs. The genomic inflation factor (lambda) was 1.0103. This value is very close to 1, suggesting that the potential inflation of test statistics due to population stratification or other systematic biases is minimal.

TS_EURO | EU | 36738982, 24927591

Children participating in the European Multicentre Tics in Children Study (EMTICS) were recruited from multiple sites in Europe and were combined with samples from the TS-EUROTRAIN study and the TSGeneSEE (Tourette Syndrome Genetics: Southern and Eastern Europe Initiative) study (Supplementary Table 21). The ascertainment of these samples was described previously in Tsetsos et al[12]. All 1,225 cases met DSM-IV-TR or DSM-5 criteria for TS or chronic motor or vocal tic disorders and were genotyped on the Illumina HumanOmniExpress BeadChip at Decode genetics. Ancestry-matched controls include 1,782 controls contributed by TS-EUROTRAIN/TSGeneSEE (949 controls from Denmark, 293 controls from Hungary, and 540 controls from Poland, Supplementary Table 20), 1,085 controls from Genomic Psychiatry Cohort (GPC), all genotyped on Illumina HumanOmniExpress; 237 controls from Greece and 38 controls from Italy genotyped on Illumina HumanHap2.5M array; and 199 Italian controls from the Hypergenes project genotyped on Illumina 1M-Duo array[13]. All cases and controls were merged into one data set, and only the overlapping SNPs across all datasets were used for GWAS. The same standard quality control and imputation steps applied on TAAICG_610K (described in session TAAICG_610K | USA/EU/CA | 22889924) were conducted on the EMTICS data set. The association test was performed on SNPs with

INFO score > 0.8 and MAF > 0.01, using a logistic regression model in PLINK2 with the first 8 and the $14^{th}$, $16^{th}$, and $19^{th}$ MDS components as covariates.

FinnGen | Finland | 36653562

The summary statistics for the tic disorder GWAS in the Finnish biobank were provided by FinnGen[14]. This dataset comprises 293 cases and 342,206 controls, with all cases meeting the criteria of ICD-10 code F95 (all forms of tic disorders), ICD-9 code 307.2 (tics), or ICD-8 code 306.20. The individuals were genotyped with Illumina and Affymetrix chip arrays (Illumina Inc., San Diego, and Thermo Fisher Scientific, Santa Clara, CA, USA). Quality control of both individuals and SNPs was conducted using the FinnGen pipeline (https://finngen.gitbook.io/documentation/v/r8/methods/finemapping). The SNPs were imputed in Beagle 4.1[15] using 3,775 Finnish whole-genome sequences (16,962,023 variants) as the reference panel. Imputed SNPs were subsequently filtered based on INFO score (>=0.6) and minimum allele counts (MACL>=5). Principal components analysis was conducted on 200,000 SNPs after LD pruning. The GWAS was executed using regenie, with age, sex, 10 PCs, genotyping batch, and the genetic relationship matrix included as the covariates[16].

iPSYCH | Denmark | 41435841

Danish nationwide population-based case-control samples were collected in the scope of the 'The Lundbeck Foundation Initiative for Integrative Psychiatric Research' (iPSYCH). Samples stem from the newly updated baseline cohort iPSYCH2015 including singletons born between 1981 and 2008 who were born to a known mother and resided in Denmark on their first birthday. Heel prick blood samples were collected from all babies and genetic information was obtained by the Statens Serum Institut (SSI) at the Danish Neonatal Screening Biobank (DNSB). The genetic information was linked with the Danish Civil Registration System and thereby coupled with the Danish Psychiatric Central Research Register which collects patient data of individuals treated in psychiatric hospitals (from 1969 onwards) or in outpatient psychiatric clinics (from 1995 onwards). General details about the cohort can be found in the primary publications[17,18].

Cases included in the present study were not primarily ascertained for TS/PTD; TS/PTD cases were drawn from cases that also presented a diagnosis of one of the core disorders iPSYCH primarily collected for and from randomly ascertained population "controls" with a diagnosis of TS/PTD. TS/PTD cases were diagnosed by a healthcare professional and met ICD-10 (F95 category: F95.0, F95.1, F95.2, F95.8 and F95.9) criteria, controls were randomly selected (down sampled to 10 times the case N) from the controls and excluded individuals with an F95 diagnosis. The final data set included 3,041 TS/PTD cases and 29,808 controls. Of the cases, 525 are female (17.26%), 23 (0.76%) are comorbid with schizophrenia (SCZ), 22 (0.72%) are comorbid with schizophrenia spectrum disorder (SCZspec), 98 (3.22%) are comorbid with affective disorders (AFF), 648 (21.31%) are comorbid with autism spectrum disorder (ASD), 1,014 (33.34%) are comorbid with attention-deficit hyperactivity disorder (ADHD), 15 (0.49%) are comorbid with OCD, and 1078 (35.45%) are comorbid with more than one of the aforementioned disorders. Of the controls, 14,683 are female (49.26%), 68 (0.23%) are diagnosed with SCZ, 60 (0.20%) are diagnosed with SCZspec, 550 (1.84%) are diagnosed with AFF, 265 (0.89%) are diagnosed with ASD, 350 (1.17%) are diagnosed with ADHD, 149 (0.50%) are diagnosed with OCD, 335 (1.12%) are diagnosed with more than one of the aforementioned disorders, and 28,031 (94.04%) are not diagnosed with any of the aforementioned disorders.

Samples for iPSYCH2012 were genotyped on the PsychChip v 1.0 array (Illumina, San Diego, CA, USA), while samples for iPSYCH2015i were genotyped on the Illumina Global Screening (GSA) v2 Array, both at the Broad Institute of MIT and Harvard (Cambridge, MA, USA). Genotype calling of markers with MAF > 0.01 was performed by merging call sets from GenCal[19] and Birdseed[20], and less frequent variants were called with zCall[21]. Genotyping and data analysis was performed in 23 waves.

Genotype data were processed using RICOPILI[22] to perform stringent QC, imputation, PC analysis, and primary association analysis. SHAPEIT was used for phasing, imputation was conducted with IMPUTE2, using the HRC as a reference panel[23]. We removed samples with a call rate below 95%, with a sex mismatch, between the sex obtained from genotype data and from the register data, as well as related individuals. PC analysis was used to exclude ancestral outliers of non-European descent, excluding all individuals exceeding eight standard deviations from the mean on the first three PCs. The association test was conducted using logistic regression model in PLINK, and the first 10 PCs were included as covariates.

MoBa | Norway | 27063603, 40834906

The Norwegian Mother, Father, and Child Cohort Study (MoBa) is a population-based cohort study conducted by the Norwegian Institute of Public Health. Participants were recruited from all over Norway from 1999-2008. The women consented to participation in 41% of the pregnancies. The cohort includes approximately 114,500 children, 95,200 mothers and 75,200 fathers. Blood samples were obtained from the mothers and fathers at 17–18 weeks of gestation and from mothers and children (umbilical cord) at birth (see https://doi.org/10.5324/nje.v24i1-2.1755 for more details).

Genotyping of MoBa has been conducted through multiple research projects, spanning several years, involving various selection criteria, and genotyping centers. Specifically, MoBa was genotyped in 26 batches and sub-batches, using three genotyping arrays: HumanCoreExome (HCE), OmniExpress (OMNI) and Global Screening Array (GSA). Phasing and imputation were performed using the publicly available European Genome-Phenome Archive (Study ID EGAS00001001710) Haplotype Reference Consortium release 1.1 as the reference panel[23]. After post-imputation QC the output of the MoBa PsychGen pipeline included 207,569 unique individuals (76,577 children, 53,358 fathers, and 77,634 mothers) of European ancestry and 6,981,748 autosomal SNPs. MoBa genotyping, quality control, phasing, imputation, and post-imputation quality control has previously been described https://doi.org/10.1101/2022.06.23.496289.

The TD cases were obtained from the MoBa offspring generation by linkage to International Classification of Diseases (ICD)-10 diagnostic information from the Norwegian Patient Registry (NPR), available from

2008 to 2021. Cases were defined by at least one contact due to either chronic motor or vocal tic disorder (F95.1) or Tourette's disorder (F95.2). This resulted in a total of 699 cases (506 boys and 193 girls). Our controls were defined as having no F95 diagnosis registered in the NPR. Due to the large case/control imbalance, we randomly selected 13,980 controls (matched by sex and genotyping batch) so that cases represented approximately 5% of the total sample size.

PLINK version 1.9 was used to run an unrelated association analysis (as there was a low amount, less than 1%, of relatedness between the cases in our sample). The first 10 PCs and genotyping batch were included as covariates.

Ethics

The establishment of MoBa and initial data collection was based on a license from the Norwegian Data Protection Agency and approval from The Regional Committees for Medical and Health Research Ethics. The MoBa cohort is now based on regulations related to the Norwegian Health Registry Act. The current study was approved by The Regional Committees for Medical and Health Research Ethics (10140).

Acknowledgements

We are grateful to all the participating families in Norway who take part in this on-going cohort study. For generating high-quality genomic data, we thank the Norwegian Institute of Public Health (NIPH), the HARVEST collaboration, the NORMENT Centre at the University of Oslo, the Center for Diabetes Research at the University of Bergen, deCODE Genetics, the Research Council of Norway, the South-Eastern and Western Norway Regional Health Authorities, the ERC AdG, Stiftelsen KG Jebsen, the Trond Mohn Foundation, and the Novo Nordisk Foundation.

Disclaimer Data from the Norwegian Patient Registry has been used in this publication. The interpretation and reporting of these data are the sole responsibility of the authors, and no endorsement by the Norwegian Patient Registry is intended nor should be inferred.

This work was performed on the TSD (Tjeneste for Sensitive Data) facilities, owned by the University of Oslo, operated and developed by the TSD service group at the University of Oslo, IT Department (USIT).. The analyses were performed on resources provided by Sigma2 - the National Infrastructure for High-Performance Computing and Data Storage in Norway.

NORDiC_SWE | Sweden | 31424634

A paper describing the rationale, design and methods of the NORDiC study has been published previously[24]. Tourette syndrome (TS) and persistent tic disorder (PTD) cases from Sweden were collected as part of the larger NORDiC study. In brief, participants were recruited from specialist clinics across Sweden, a clinical trial[25], or self-referral via a dedicated study website. Diagnoses were confirmed by a diagnostic interview in all cases. Participants were children or adults with a diagnosis of either TS or PTD. Case samples were genotyped on an Illumina Global Screening Array (GSA) v3 platform in Bonn, Germany by LIFE&BRAIN. Swedish controls were collected as part of LifeGene[10], a prospective population-based cohort of around 50,000 individuals in Sweden. We obtained a dataset of 500 LifeGene controls genotyped on a GSA v2 array.

TAAICG_610K | USA/EU/CA | 22889924

Cases of TAAICG_610K were ascertained from multiple sites in the USA, Canada, UK, Netherlands, and Israel by the Tourette Association of America International Consortium for Genetics (TAAICG). All cases were defined as TS by the TS Classification Study Group (TSCSG) according to DSM-IV-TR criteria plus tics observed by an experienced clinician[26]. Controls were recruited from the Center for Applied Genomics (CAG) at the Children's Hospital of Philadelphia (CHOP) (N = 776) and from the Prostate Cancer Study by the Breast and Prostate Cancer Cohort Consortium (BPC3) (N = 1,247)[27]. All samples were genotyped on the Illumina Human610-Quadv1_B SNP array (Illumina, San Diego, CA, USA). Standard quality control (QC) protocol was conducted with PLINK[28].

Standard quality control steps were conducted on TAAICG_610K data. Samples were removed for call rates < 98%, sex discrepancy, ambiguous genomic sex, inbreeding coefficient |F|>0.2, or related samples with pi_hat > 0.2. SNP QC included removing monomorphic SNPs, CNV-targeted SNP probes, SNPs with genotyping rate < 98%, SNPs with minor allele frequency (MAF) < 0.01, strand-ambiguous SNPs with significant allele frequency differences or aberrant LD correlations with adjacent SNPs based on the entire HapMap2 reference panel, SNPs with $P < 1.0 \times 10^{-6}$ in Hardy Weinberg Equilibrium (HWE) test among controls or $P < 1.0 \times 10^{-10}$ among cases, SNPs with differential missing rate between cases and controls (> 0.02), and SNPs with batch effect ($P < 1.0 \times 10^{-5}$) between two control cohorts. Multidimensional scaling (MDS) analyses were performed in PLINK2 and samples were removed when they were significant outliers in the first five MDS dimensions or when there were no matching cases or controls on these MDS dimensions. In addition, the samples that were related to any samples in other studies were removed from this data. The genotyped data were phased by SHAPEIT2[29] and imputed by Minimac3[30] using the HRC 24 release 1.1 as the reference panel. The association test was conducted on SNPs with an INFO score > 0.8 and MAF > 0.01, using a logistic regression model in PLINK2 with the first five MDS components as covariates. Additionally, three MDS components (7$^{th}$, 13$^{th}$, and 17$^{th}$) were found significantly associated with TS status and were included in the logistic regression as covariates.

TAAICG_Fam | USA/EU/CA | 30818990

Cases in TAAICG_Fam came from the subjects who were recruited by TAAICG and had at least one related family member diagnosed with DSM-IV or DSM-5 Tourette's syndrome (N=818). The ancestry-matched controls came from two studies: 1) 812 controls were from Late-Onset Alzheimer's Disease and National Cell Repository for Alzheimer's Disease Family Study[31], and 2) 950 controls were the controls recruited by the National Institute of Neurological Disorders and Stroke (NINDS). All subjects were genotyped on the Illumina Human610-Quadv1_B SNP array or HumanOmniExpressExome_8v1 array. The standard quality control steps were conducted on the SNPs genotyped on both arrays using unrelated subjects (i.e. one subject per sample). The sample quality control included removing subjects with call rates < 98%, sex discrepancy or ambiguous genomic sex, or inbreeding coefficients |F|>0.2. Non-European ancestry samples were identified by FRAPOSA[32] and excluded. The same imputation process as in TAAICG_610K was conducted, and the association test was performed on the SNPs with INFO>0.8 and MAF>0.01 using a linear mixed model in MMM[33] to account for the relatedness among cases.

TAAICG_GSA | USA

Cases in TAAICG_GSA were recruited from Tourette's syndrome specialty clinics in the United States (N = 209) by TAAICG, and all cases were diagnosed with DSM-5 Tourette's syndrome after a structured interview. Eighteen ancestry-matched controls were recruited together with the cases. Additional ancestry-matched controls came from Genomic Psychiatry Cohort (N = 440)[34]. All samples were genotyped on the Infinium Global Screening Array (GSAMD-24v1-0_20011747_A1) bead chip (Illumina). The same standard quality control and imputation steps applied on TAAICG_610K were conducted on the TAAICG_GSA data set. The association test was performed on SNPs with an INFO score > 0.8 and MAF > 0.01, using a logistic regression model in PLINK2 with the first four MDS components as covariates.

TAAICG_Omni | USA/EU/CA | 30818990

Cases in TAAICG_Omni were recruited by TAAICG from two sources: 1) 1,280 cases with DSM-5 Tourette's syndrome were recruited by e-mail or online recruitment combined with validated, web-based phenotypic assessments[35,36]; 2) 1,468 cases were recruited from Tourette's syndrome specialty clinics in the United States, Canada, the Netherlands, Austria, France, Germany, Greece, Hungary, and Italy. Subjects with a history of intellectual disability or with tics or movement disorder phenocopies caused by genetic or neurological disorders were excluded. OCD and ADHD were phenotypically assessed by clinicians with expertise in the diagnosis of Tourette syndrome, OCD, and ADHD, and diagnoses were assigned based either on DSM-5 criteria alone or by using standardized structured instruments[35,36]. Ancestry-matched controls came from five studies: 1) 486 controls were collected from Tourette's syndrome specialty clinics along with the cases; 2) 640 controls came from Consortium for Neuropsychiatric Phenomics (CNP) Controls[37]; 3) 924 controls were from Genomics Superstruct Project (GSP)[38]; 4) 856 controls were the Ashkenazi Jewish controls from the Center for Inherited Disease Research (CIDR)[39]; 5) 231 Italian controls came from the Hypergenes project[13]. The Italian controls were genotyped on the Illumina 1M-Duo chip, and the remaining cases and controls were genotyped on the Illumina HumanOmniExpressExome_8v1 array (Illumina, San Diego, CA). The same standard quality control and imputation steps applied on TAAICG_610K were conducted on the TAAICG_Omni data set. GWAS was performed on SNPs with INFO score > 0.8 and MAF > 0.01, using a logistic regression model in PLINK2 with the first six and the 19th MDS components as covariates.

TAAICG_PsychChip | USA

Cases and controls in TAAICG_PsychChip were recruited from Tourette's syndrome specialty clinics in the United States by TAAICG and all cases were diagnosed with DSM-5 Tourette's syndrome after structured interview. Additional ancestry-matched controls were selected from Mass General Brigham controls. All samples were genotyped on the Illumina PsychChip _15048346_B array. The same standard quality control and imputation steps applied on TAAICG_610K were conducted on the TAAICG_PsychChip data set. After quality control steps, cross-dataset IBD was estimated to identify samples with pi_hat>0.2 with samples in another data set, and these samples were excluded from TAAICG_PsychChip. The remaining samples contain 36 cases and 178 controls. The association test was performed on SNPs with INFO score > 0.8 and MAF > 0.01, using a logistic regression model in PLINK2 with the first four and the 17th MDS components as covariates.

TIC_Genetics | USA | 30818990

Cases participating in the Tourette International Collaborative Genetics study (TIC_Genetics) were recruited as probands of parent-child trios. The ascertainment of these samples was described previously in Dietrich et al[40]. All 532 cases meet the criteria for DSM-IV-TR TS or chronic motor or vocal tic disorder. The cases were genotyped on three different Illumina platforms: 347 cases were genotyped on Illumina HumanOmniExpress array, 107 cases were genotyped on the Illumina HumanHap1M array, and 78 cases were genotyped on the Illumina HumanHap2.5M array. Ancestry-matched controls include 1,285 controls from the Studies of Addiction: Genetics and Environment (SAGE) genotyped on Illumina HumanHap1Mv1_C array and 163 Ashkenazi Jewish controls from CIDR genotyped on Illumina HumanOmniExpressExome_8v1 array[39]. All cases and controls were merged into one data set and only the overlapping SNPs across all datasets were used for GWAS. The same standard quality control and imputation steps applied on TAAICG_610K were conducted on the TIC_Genetics data set. The association test was performed on SNPs with INFO score > 0.8 and MAF > 0.01, using a logistic regression model in PLINK2 with the first 3 and the 11$^{th}$ and 20$^{th}$ MDS components as covariates.

Before the association test in each data set, the identity by descent (IBD) was estimated across all data sets with available individual-level genotyping data. For any cross-dataset duplicates or related pairs, the sample from a less informative genotyping platform was removed from this GWAS.

**Supplementary Notes 3: Gene-based Tests**

We carried out a series of gene-based tests using MAGMA v1.10 to determine if any annotated genes carry a significant amount of TS risk variation in the summary statistics. We applied these tests to a high-quality subset of the TS GWAS summary statistics. To derive these summary statistics, we took a subset of the SNPs that carried a MAF>0.01 and INFO>0.6. The summary statistics utilized for gene-based tests included a total of 6019193 SNPs.

Gene annotations for a total of 19,175 gene units were taken from Bryois et al.[41] and can be found at https://github.com/jbryois/scRNA_disease/raw/refs/heads/master/Code_Paper/Data/NCBI/NCBI37.3.gene.loc.extendedMHCexcluded. We focused our analyses on the subset of these gene units corresponding to protein-coding genes from Ensembl version 87. Three separate tests were conducted: standard MAGMA, testing individual gene units defined by an interval 35 kilobases upstream of the transcription start site to 10 kilobases downstream of the transcription stop site (18,028 tests); e-MAGMA, where SNPs are defined as members of a gene if they were defined as eQTLs in GTEx in at least one brain tissue sampled (9446 tests); h-MAGMA, where loci for a given gene include both the main gene body from MAGMA and distal intronic/intergenic regions overlapping a chromatin interaction detected in fetal or adult brain that linked back to the gene body (18,364 tests). All MAGMA tests used the 'effective sample size', defined as 4*Nca*Nco/(2*(Nca+Nco)), as the total sample size per SNP. In total, we conducted 45,838 gene-based tests using the TS summary statistics, and defined significance based on a Bonferroni threshold of $0.05/45,838 = 1.1 \times 10^{-6}$.

Of the 45,838 tests conducted, a total of 29 tests passed the Bonferroni threshold for significance. Of these 29 tests, 13 were MAGMA-based, 3 were from E-MAGMA, and 13 were from H-MAGMA. These tests highlight a total of 25 distinct protein-coding genes. Only one protein-coding gene, *WDR6*, is significant in all 3 test sets.

**Supplementary Notes 4: Tissue/cell-type Enrichment Analyses**

For our tissue and cell-type enrichment analyses, we utilized a protocol defined and described within[41]. Consistent with this, sets of genes that are defined as marking specific tissues and cell types are derived from this publication, and can be extracted from https://github.com/jbryois/scRNA_disease.

We included 3 datasets in our analysis, which had been preprocessed and made available by Bryois et al[41].The first comes from GTEx[42] and includes a total of 37 tissues that are testable. The other 2 datasets come from Zeisel et al[43] and represent broad and specific neuronal cell types (39 and 265 testable cell types, respectively) derived from single-cell RNA sequencing of a whole mouse brain.

For each of the 3 datasets, we applied the protocol described in Bryois et al[41]. In general, the protocol for a given set of genes representing a tissue or cell type involves applying both a MAGMA geneset-based test and an LDSC partitioned heritability test using SNPs that fall within the standard MAGMA gene locus (35 kb upstream to 10 kb downstream). For a given set of tests, a tissue or cell type were only considered significant if FDR-adjusted p-values from both MAGMA and LDSC are less than 0.05. Since the LDSC tests were significantly more computationally intensive than MAGMA tests, for tests of specific cell types from Zeisel et al[43] (265 cell types total) we only performed tests using MAGMA and considered a specific neuronal cell type as significant if the FDR-adjusted p-value is less than 0.05. Finally, we derived cell-type 'clusters' based on the 'Description' column from the Zeisel et al. level 5 cell-type table and only retained those that had a membership of 2 or more (35 total), and defined a single mean $-\log_{10}(P)$ per cluster using the specific cell-type statistics. We applied FDR-adjustment to these cluster-level p-values and labelled clusters as significant if FDR-adjusted p-values were <0.05.

Statistical analysis and plotting done downstream of MAGMA and LDSC were conducted using R v4.2.1. In gene-based tests, Bonferroni correction for multiple testing was conducted using the function p.adjust (method="Bonferroni"). In tissue and cell-type analyses, FDR correction for multiple testing was performed using the function p.adjust(method="fdr"). For all plotting we utilized the R package ggplot2 v3.4.4.

### Supplementary Figures

A.

B.

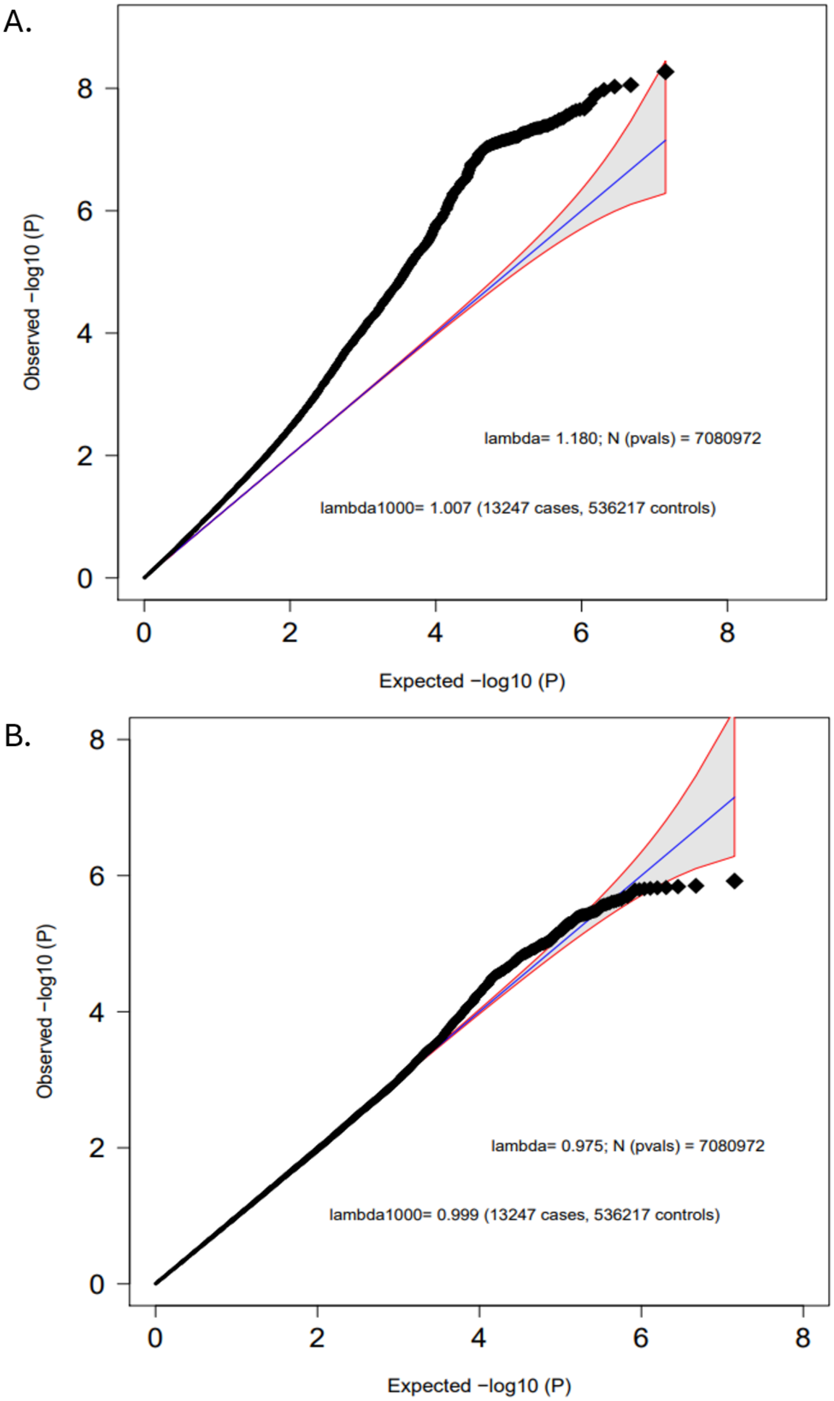


**Supplementary Figure 1**. A) Quantile-Quantile (Q-Q) plot of observed versus expected association -$\log_{10}$(*P*-values) for the TD meta-analysis GWAS, including 13,247 cases and 536,217 controls. The genomic inflation factor (lambda) is 1.007 after scaling to 1000 cases and 1000 controls. B) Q-Q plot of observed versus expected **heterogeneity** -$\log_{10}$(*P*-values) for the TD meta-analysis GWAS. The genomic inflation factor (lambda) is 0.999 after scaling to 1000 cases and 1000 controls.

A.

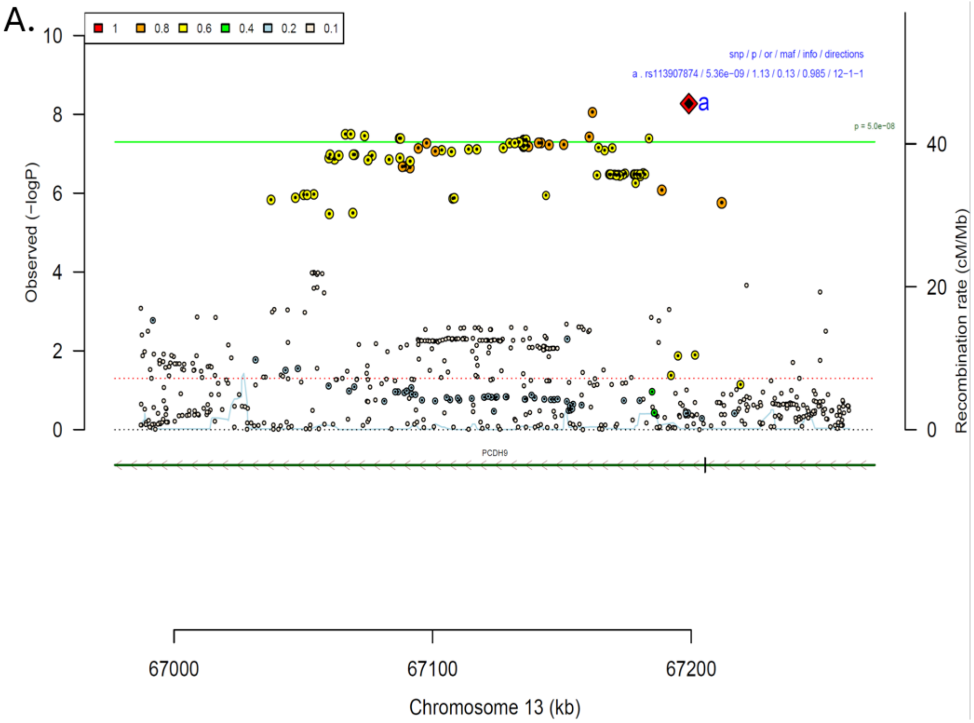


B.

| rs113907874 | | T/C | | 13:67199085 | | | |
|---|---|---|---|---|---|---|---|
| | | +++++?−+++++++ | | het_P: | 0.308 | het_I: | 13.6 |

| | ngt | info | p_value | f_ca(n) | f_co(n) | ln(OR) | STDerr |
|---|---|---|---|---|---|---|---|
| dan_TSCT_deCODE_AFco0.01.assoc. | 0 | 1.00 | 0.00225 | 0.127(1998) | 0.127(134508) | 0.163 | 0.0533 |
| dan_TS_610k_INFO0.8_AFco0.01.assoc. | 0 | 0.92 | 0.237 | 0.148(857) | 0.138(2256) | 0.105 | 0.089 |
| dan_TS_BioVU_AFco0.01.assoc. | 1 | 1.00 | 0.877 | 0.141(319) | 0.140(1585) | 0.0198 | 0.129 |
| dan_TS_EMTICS_noSpanish_AFco0.01.assoc. | 0 | 0.97 | 0.593 | 0.169(1225) | 0.162(3341) | 0.035 | 0.0655 |
| dan_TS_Fam_AFco0.01.assoc. | 0 | 0.89 | 0.303 | 0.143(818) | 0.138(1762) | 0.0733 | 0.0712 |
| dan_TS_GSA_INFO0.8_AFco0.01.assoc. | 0 | 0.92 | 0.51 | 0.138(209) | 0.152(458) | −0.118 | 0.18 |
| dan_TS_Moba_INFO0.8_AFco0.01.assoc. | 0 | 0.97 | 7.64e−05 | 0.178(699) | 0.139(13980) | 0.289 | 0.0731 |
| dan_TS_NORDiCSWE_AFco0.01.assoc. | 0 | 0.97 | 0.525 | 0.175(188) | 0.160(414) | 0.109 | 0.172 |
| dan_TS_Psych_AFco0.01.assoc. | 0 | 0.96 | 0.159 | 0.201(36) | 0.128(178) | 0.516 | 0.367 |
| dan_TS_Sweden.assoc. | 0 | 0.98 | 0.416 | 0.169(284) | 0.157(1136) | 0.103 | 0.126 |
| dan_TS_TAAICG_woEGA_filtered.assoc. | 0 | 0.96 | 0.397 | 0.158(2748) | 0.156(3137) | 0.0463 | 0.0547 |
| dan_TS_TICGEN_AFco0.01_2.assoc. | 0 | 0.99 | 0.266 | 0.154(532) | 0.142(1448) | 0.114 | 0.102 |
| dan_TS_iPSYCH_maf0.01_info0.8.assoc. | 0 | 0.94 | 0.000121 | 0.171(3041) | 0.152(29808) | 0.142 | 0.0371 |
| **dan_** | **1** | **0.98** | **5.36e−09** | **0.157(13247)** | **0.134(536217)** | **0.118** | **0.0202** |

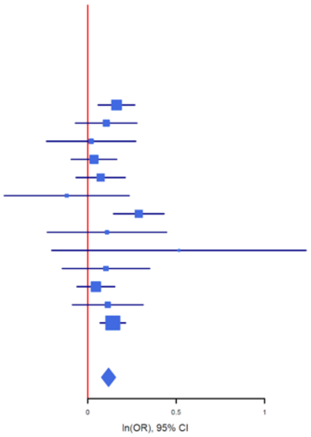


**Supplementary Figure 2**: A) **Regional association plot** surrounding SNP rs113907874 in the TD meta-analysis GWAS. The y-axis on the left shows the –log$_{10}$*P*-values of the SNP associations, while the y-axis on the right represents recombination rates (blue line) in centimorgans (cM) per megabase (Mb). The x-axis indicates the genomic position (Mb), with annotated genes displayed below the plot. The most strongly associated SNP (index SNP) is represented by a diamond and labeled with "a". The colors of the surrounding SNPs reflect their linkage disequilibrium (LD) with the index SNP. B) **Forest plot** displaying the effect sizes of rs113907874 across individual studies and the meta-analysis. The table on the left provides details on imputation quality (INFO) score, SNP association *P*-value, allele frequencies in cases (f_ca) with case sample size (n), allele frequencies in controls (f_co) with control sample size (n), beta estimates (ln(*OR*)), and standard error (STDerr) for each study and the overall meta-analysis (dan_). The forest plot visualizes the effect sizes (ln(*OR*)) and their 95% confidence intervals for each cohort and the combined meta-analysis.

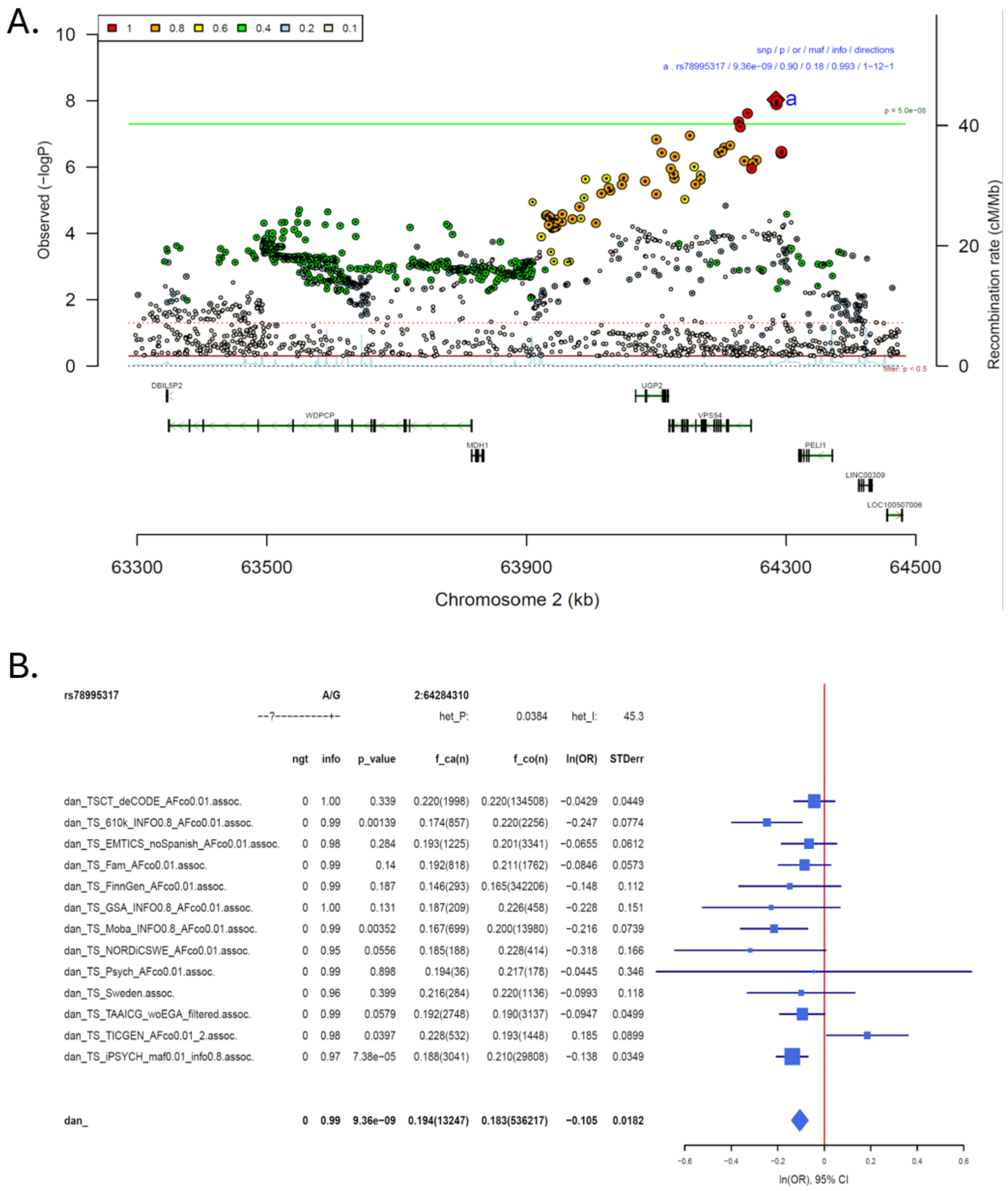


**Supplementary Figure 3**: A) **Regional association plot** surrounding SNP rs78995317 in the TD meta-analysis GWAS. The y-axis on the left shows the $-\log_{10}P$-values of the SNP associations, while the y-axis on the right represents recombination rates (blue line) in centimorgans (cM) per megabase (Mb). The x-axis indicates the genomic position (Mb), with annotated genes displayed below the plot. The most strongly associated SNP (index SNP) is represented by a diamond and labeled with “a”. The colors of the surrounding SNPs reflect their linkage disequilibrium (LD) with the index SNP. B) **Forest plot** displaying the effect sizes of rs113907874 across individual studies and the meta-analysis. The table on the left provides details on imputation quality (INFO) score, SNP association *P*-value, allele frequencies in cases (f_ca) with case sample size (n), allele frequencies in controls (f_co) with control sample size (n), beta estimates (ln(*OR*)), and standard error (STDerr) for each study and the overall meta-analysis (dan_). The forest plot visualizes the effect sizes (ln(*OR*)) and their 95% confidence intervals for each cohort and the combined meta-analysis.

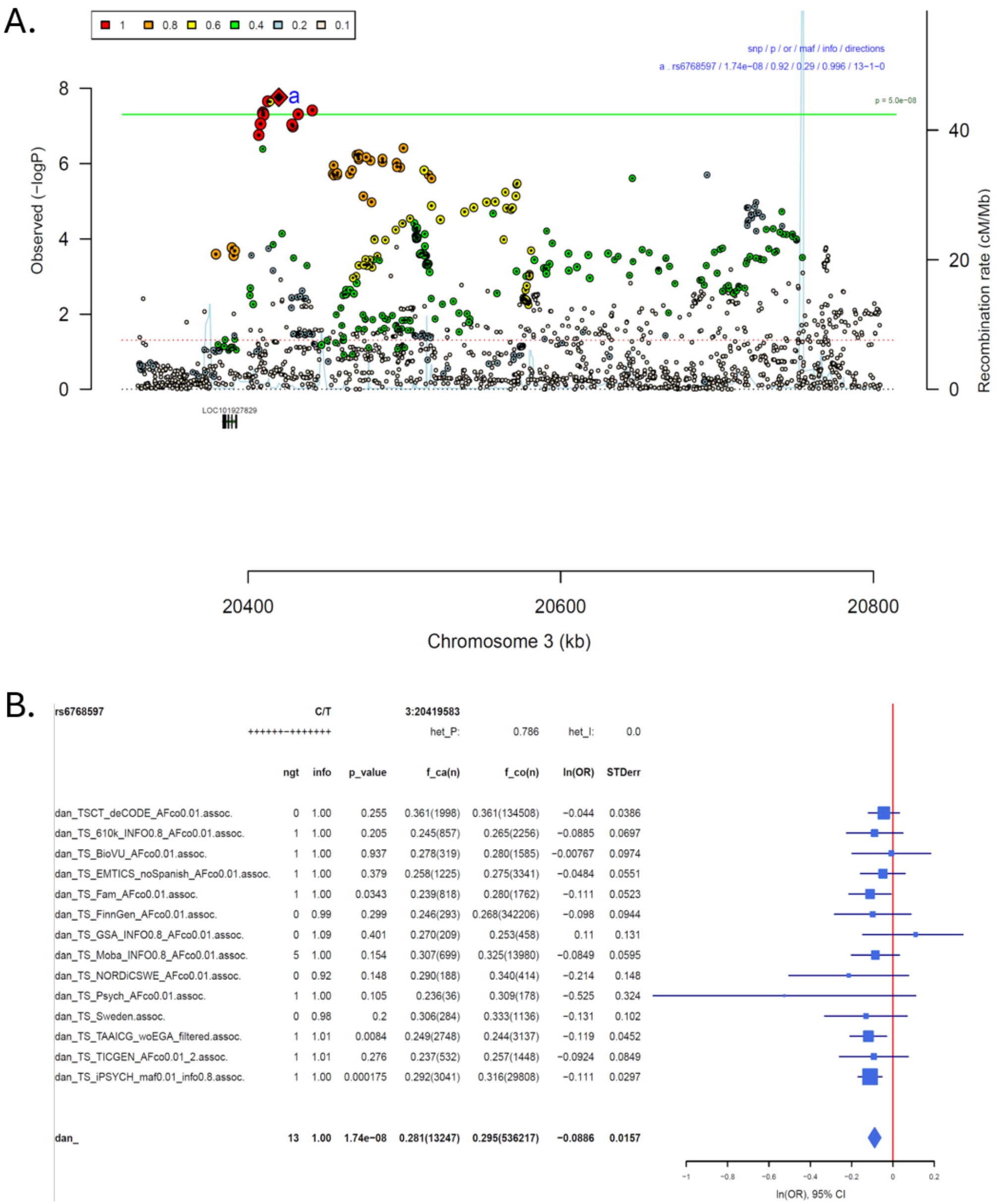


**Supplementary Figure 4**: A) **Regional association plot** surrounding SNP rs6768597 in the TD meta-analysis GWAS. The y-axis on the left shows the $-\log_{10}P$-values of the SNP associations, while the y-axis on the right represents recombination rates (blue line) in centimorgans (cM) per megabase (Mb). The x-axis indicates the genomic position (Mb), with annotated genes displayed below the plot. The most strongly associated SNP (index SNP) is represented by a diamond and labeled with “a”. The colors of the surrounding SNPs reflect their linkage disequilibrium (LD) with the index SNP. B) **Forest plot** displaying the effect sizes of rs113907874 across individual studies and the meta-analysis. The table on the left provides details on imputation quality (INFO) score, SNP association *P*-value, allele frequencies in cases (f_ca) with case sample size (n), allele frequencies in controls (f_co) with control sample size (n), beta estimates (ln(*OR*)), and standard error (STDerr) for each study and the overall meta-analysis (dan_). The forest plot visualizes the effect sizes (ln(*OR*)) and their 95% confidence intervals for each cohort and the combined meta-analysis.

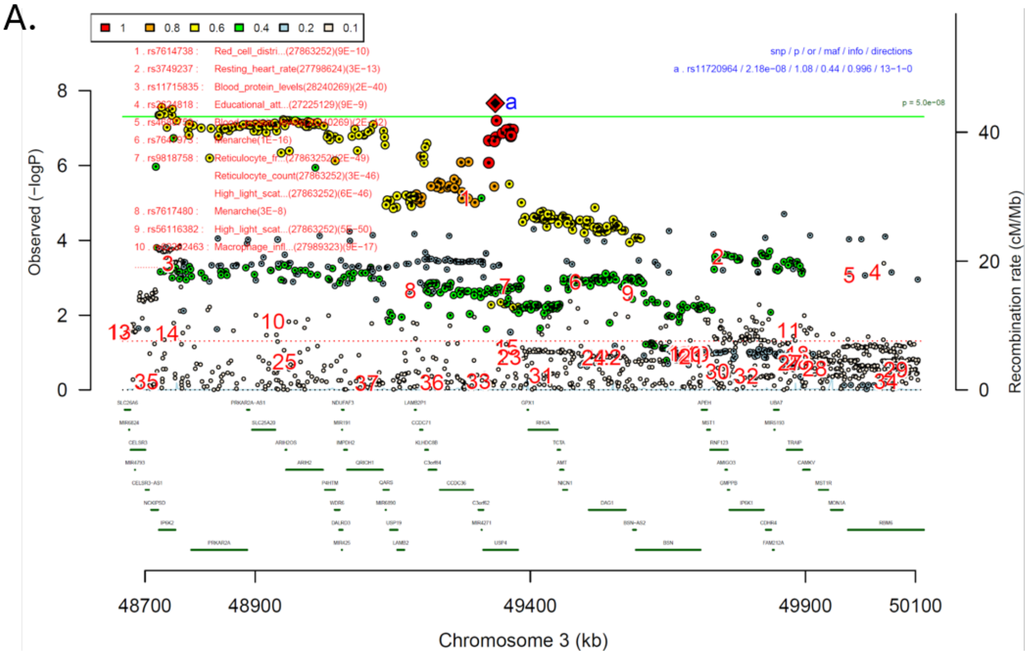


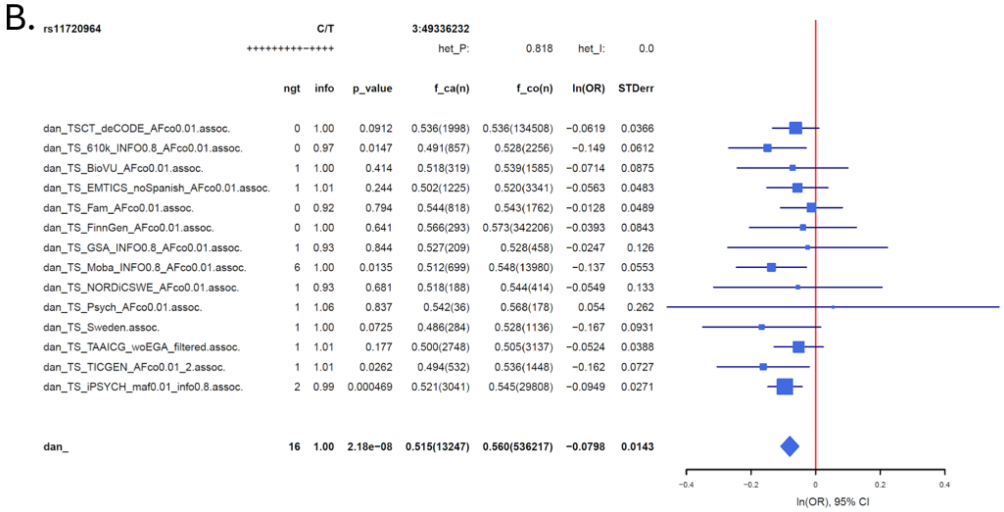


**Supplementary Figure 5**: A) **Regional association plot** surrounding SNP rs11720964 in the TD meta-analysis GWAS. The y-axis on the left shows the –$\log_{10}P$-values of the SNP associations, while the y-axis on the right represents recombination rates (blue line) in centimorgans (cM) per megabase (Mb). The x-axis indicates the genomic position (Mb), with annotated genes displayed below the plot. The most strongly associated SNP (index SNP) is represented by a diamond and labeled with "a". The colors of the surrounding SNPs reflect their linkage disequilibrium (LD) with the index SNP. B) **Forest plot** displaying the effect sizes of rs113907874 across individual studies and the meta-analysis. The table on the left provides details on imputation quality (INFO) score, SNP association *P*-value, allele frequencies in cases (f_ca) with case sample size (n), allele frequencies in controls (f_co) with control sample size (n), beta estimates (ln(*OR*)), and standard error (STDerr) for each study and the overall meta-analysis (dan_). The forest plot visualizes the effect sizes (ln(*OR*)) and their 95% confidence intervals for each cohort and the combined meta-analysis.

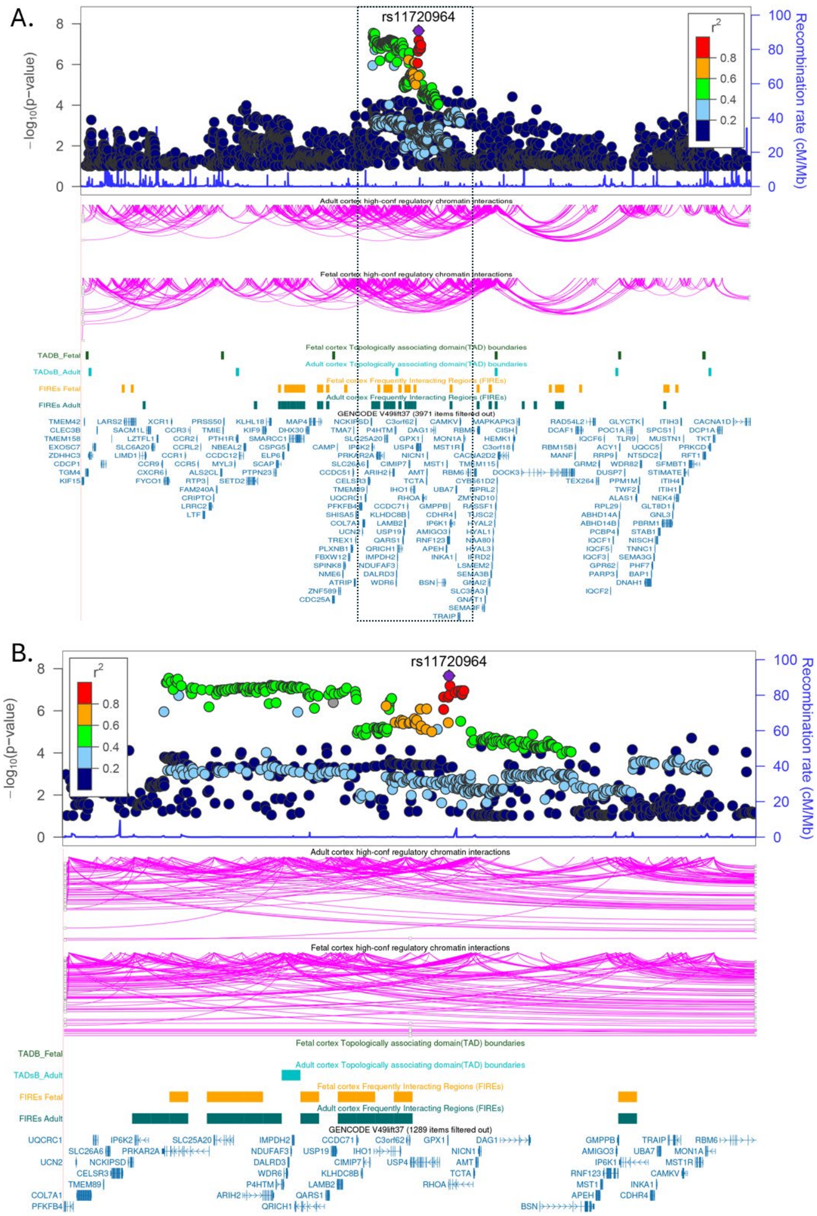


**Supplementary Figure 6**: **Functional genomic architecture of the tic disorder (TD) genome-wide significant locus at chromosome 3p21.** A) Regional Overview of the 3p21 Locus. The top panel presents a locus plot displaying -$\log_{10}$(P) values for association with TD. SNPs are color-coded based on their linkage disequilibrium ($r^2$) with the lead index SNP, rs11720964 (indicated by the purple diamond). The light blue line represents the local recombination rate in cM/Mb. Below the locus plot, high-confidence regulatory chromatin interactions are mapped for both adult cortex and fetal cortex. These are followed by structural annotations, including Topologically Associating Domain (TAD) boundaries for fetal and adult cortex, and Frequently Interacting Regions (FIREs) identified in fetal and adult cortical tissues. The bottom section lists genes within the region based on GENCODE V49lift37. B) Detailed View of the Associated Region. This panel provides a magnified view of the specific region demarcated by the dotted box in 6A. It offers higher resolution for the association signals, chromatin loops, and architectural features surrounding the index SNP rs11720964.

A.

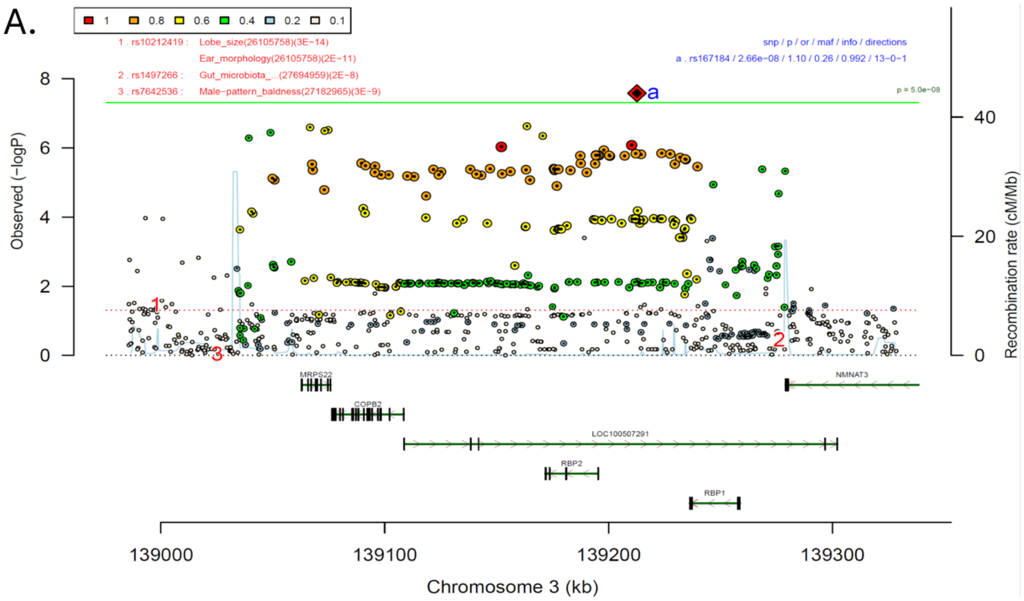


B.

| rs167184 | | | | T/C | 3:139212649 | | | |
|---|---|---|---|---|---|---|---|---|
| | | | | ++?+++++++++++ | het_P: | 0.619 | het_I: | 0.0 |
| | ngt | info | p_value | | f_ca(n) | f_co(n) | ln(OR) | STDerr |
| dan_TSCT_deCODE_AFco0.01.assoc. | 0 | 1.00 | 0.00155 | | 0.279(1998) | 0.279(134508) | 0.128 | 0.0406 |
| dan_TS_610k_INFO0.8_AFco0.01.assoc. | 0 | 0.99 | 0.0899 | | 0.291(857) | 0.261(2256) | 0.114 | 0.0673 |
| dan_TS_EMTICS_noSpanish_AFco0.01.assoc. | 0 | 0.97 | 0.497 | | 0.267(1225) | 0.261(3341) | 0.0377 | 0.0555 |
| dan_TS_Fam_AFco0.01.assoc. | 0 | 0.96 | 0.00103 | | 0.317(818) | 0.249(1762) | 0.171 | 0.052 |
| dan_TS_FinnGen_AFco0.01.assoc. | 0 | 0.99 | 0.656 | | 0.256(293) | 0.250(342206) | 0.0428 | 0.0961 |
| dan_TS_GSA_INFO0.8_AFco0.01.assoc. | 0 | 1.00 | 0.612 | | 0.288(209) | 0.270(458) | 0.0676 | 0.133 |
| dan_TS_Moba_INFO0.8_AFco0.01.assoc. | 0 | 0.98 | 0.0194 | | 0.293(699) | 0.265(13980) | 0.143 | 0.061 |
| dan_TS_NORDiCSWE_AFco0.01.assoc. | 0 | 0.96 | 0.687 | | 0.271(188) | 0.267(414) | 0.059 | 0.146 |
| dan_TS_Psych_AFco0.01.assoc. | 0 | 0.97 | 0.455 | | 0.303(36) | 0.259(178) | 0.231 | 0.309 |
| dan_TS_Sweden.assoc. | 0 | 0.97 | 0.328 | | 0.290(284) | 0.273(1136) | 0.102 | 0.104 |
| dan_TS_TAAICG_woEGA_filtered.assoc. | 0 | 0.91 | 0.533 | | 0.268(2748) | 0.281(3137) | 0.0288 | 0.0462 |
| dan_TS_TICGEN_AFco0.01_2.assoc. | 0 | 0.95 | 0.0203 | | 0.298(532) | 0.263(1448) | 0.192 | 0.0828 |
| dan_TS_iPSYCH_maf0.01_info0.8.assoc. | 0 | 0.95 | 0.0652 | | 0.269(3041) | 0.258(29808) | 0.0575 | 0.0312 |
| **dan_** | **0** | **0.99** | **2.66e−08** | | **0.278(13247)** | **0.258(536217)** | **0.0909** | **0.0163** |

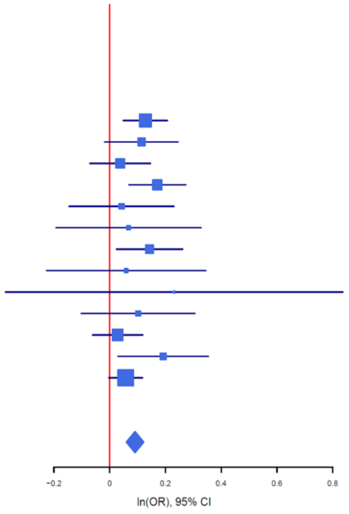


**Supplementary Figure 7**: A) **Regional association plot** surrounding SNP rs167184 in the TD meta-analysis GWAS. The y-axis on the left shows the –log$_{10}$*P*-values of the SNP associations, while the y-axis on the right represents recombination rates (blue line) in centimorgans (cM) per megabase (Mb). The x-axis indicates the genomic position (Mb), with annotated genes displayed below the plot. The most strongly associated SNP (index SNP) is represented by a diamond and labeled with "a". The colors of the surrounding SNPs reflect their linkage disequilibrium (LD) with the index SNP. B) **Forest plot** displaying the effect sizes of rs113907874 across individual studies and the meta-analysis. The table on the left provides details on imputation quality (INFO) score, SNP association *P*-value, allele frequencies in cases (f_ca) with case sample size (n), allele frequencies in controls (f_co) with control sample size (n), beta estimates (ln(*OR*)), and standard error (STDerr) for each study and the overall meta-analysis (dan_). The forest plot visualizes the effect sizes (ln(*OR*)) and their 95% confidence intervals for each cohort and the combined meta-analysis.

A

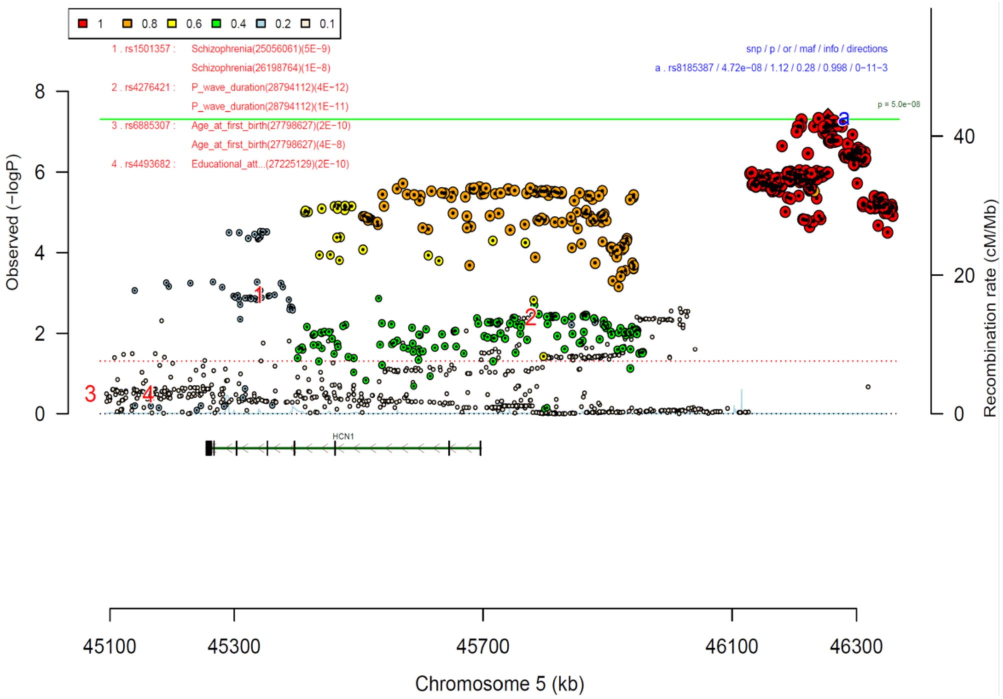


B.

| rs8185387 | | C/A | | 5:46254046 | | | |
|---|---|---|---|---|---|---|---|
|  |  | ----?-------?? |  | het_P: | 0.601 | het_I: | 0.0 |
| | ngt | info | p_value | f_ca(n) | f_co(n) | ln(OR) | STDerr |
| dan_TSCT_deCODE_AFco0.01.assoc. | 0 | 1.00 | 0.0365 | 0.311(1998) | 0.311(134508) | 0.0825 | 0.0395 |
| dan_TS_610k_INFO0.8_AFco0.01.assoc. | 0 | 0.96 | 0.00591 | 0.344(857) | 0.310(2256) | 0.18 | 0.0654 |
| dan_TS_BioVU_AFco0.01.assoc. | 0 | 1.00 | 0.253 | 0.350(319) | 0.326(1585) | 0.106 | 0.0931 |
| dan_TS_EMTICS_noSpanish_AFco0.01.assoc. | 0 | 1.00 | 0.0952 | 0.310(1225) | 0.296(3341) | 0.0875 | 0.0524 |
| dan_TS_FinnGen_AFco0.01.assoc. | 0 | 1.00 | 0.000948 | 0.323(293) | 0.266(342206) | 0.289 | 0.0873 |
| dan_TS_GSA_INFO0.8_AFco0.01.assoc. | 0 | 1.00 | 0.336 | 0.344(209) | 0.315(458) | 0.122 | 0.127 |
| dan_TS_Moba_INFO0.8_AFco0.01.assoc. | 0 | 1.00 | 0.443 | 0.320(699) | 0.310(13980) | 0.0454 | 0.0591 |
| dan_TS_NORDiCSWE_AFco0.01.assoc. | 0 | 0.87 | 0.0974 | 0.350(188) | 0.304(414) | 0.244 | 0.147 |
| dan_TS_Psych_AFco0.01.assoc. | 0 | 1.12 | 0.994 | 0.333(36) | 0.343(178) | 0.002 | 0.275 |
| dan_TS_Sweden.assoc. | 0 | 0.95 | 0.448 | 0.314(284) | 0.301(1136) | 0.0788 | 0.104 |
| dan_TS_TAAICG_woEGA_filtered.assoc. | 0 | 0.86 | 0.012 | 0.322(2748) | 0.301(3137) | 0.114 | 0.0453 |
| **dan_** | **0** | **1.00** | **4.72e−08** | **0.322(13247)** | **0.280(536217)** | **0.11** | **0.0201** |

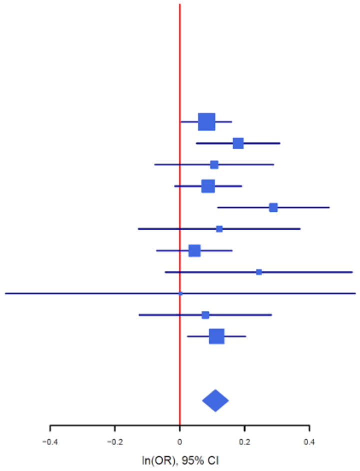


**Supplementary Figure 8**: A) **Regional association plot** surrounding SNP rs8185387 in the TD meta-analysis GWAS. The y-axis on the left shows the $–\log_{10}P$-values of the SNP associations, while the y-axis on the right represents recombination rates (blue line) in centimorgans (cM) per megabase (Mb). The x-axis indicates the genomic position (Mb), with annotated genes displayed below the plot. The most strongly associated SNP (index SNP) is represented by a diamond and labeled with "a". The colors of the surrounding SNPs reflect their linkage disequilibrium (LD) with the index SNP. B) **Forest plot** displaying the effect sizes of rs113907874 across individual studies and the meta-analysis. The table on the left provides details on imputation quality (INFO) score, SNP association *P*-value, allele frequencies in cases (f_ca) with case sample size (n), allele frequencies in controls (f_co) with control sample size (n), beta estimates (ln(*OR*)), and standard error (STDerr) for each study and the overall meta-analysis (dan_). The forest plot visualizes the effect sizes (ln(*OR*)) and their 95% confidence intervals for each cohort and the combined meta-analysis.

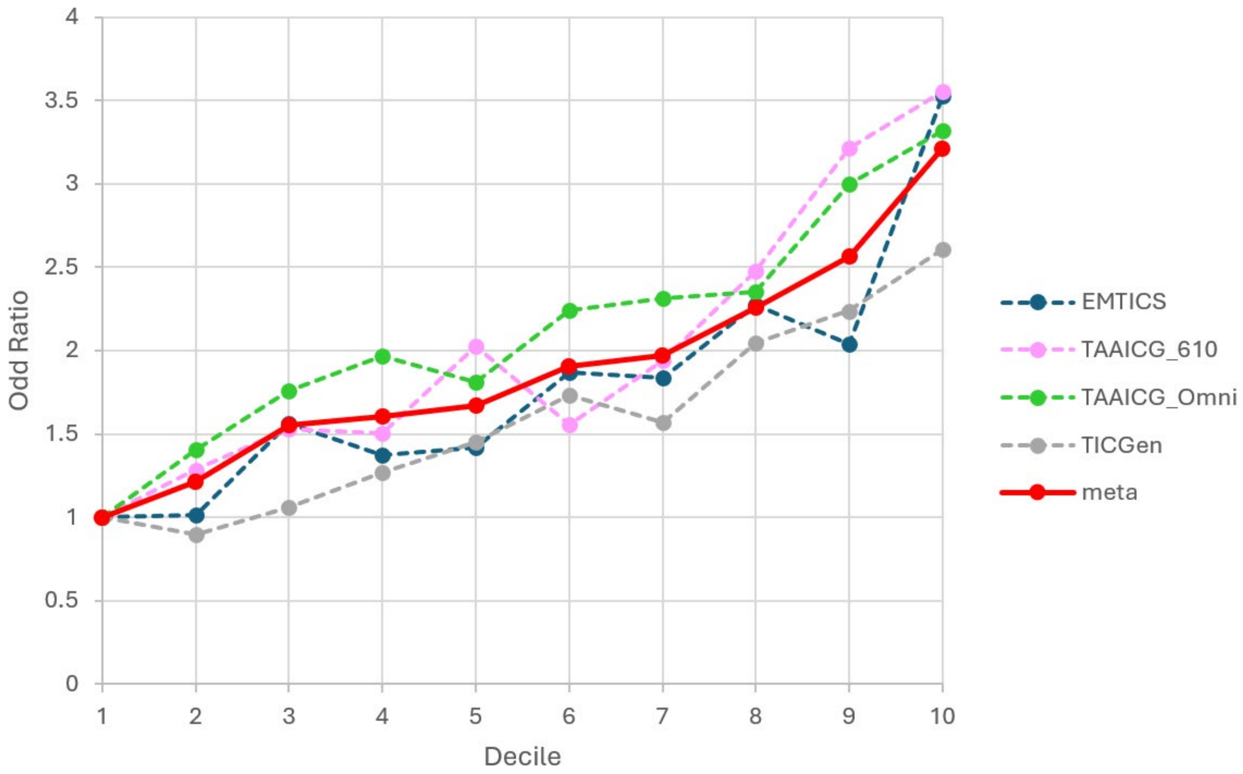


**Supplementary Figure 9**: Decile plot of odds ratios by TD PRS within each decile for four cohorts and their meta-results. The four cohorts include EMTICS (1,225 cases and 3,341 controls, blue dotted line), TAAICG_610K (857 cases and 2,256 controls, pink dotted line), TAAICG_Omni (2,748 cases and 3,137 controls, green dotted line), and TIC_Genetics (532 cases and 1,448 controls, grey dotted line). The meta-analysis of four cohorts contains 5,362 cases and 10,182 controls (red solid line).

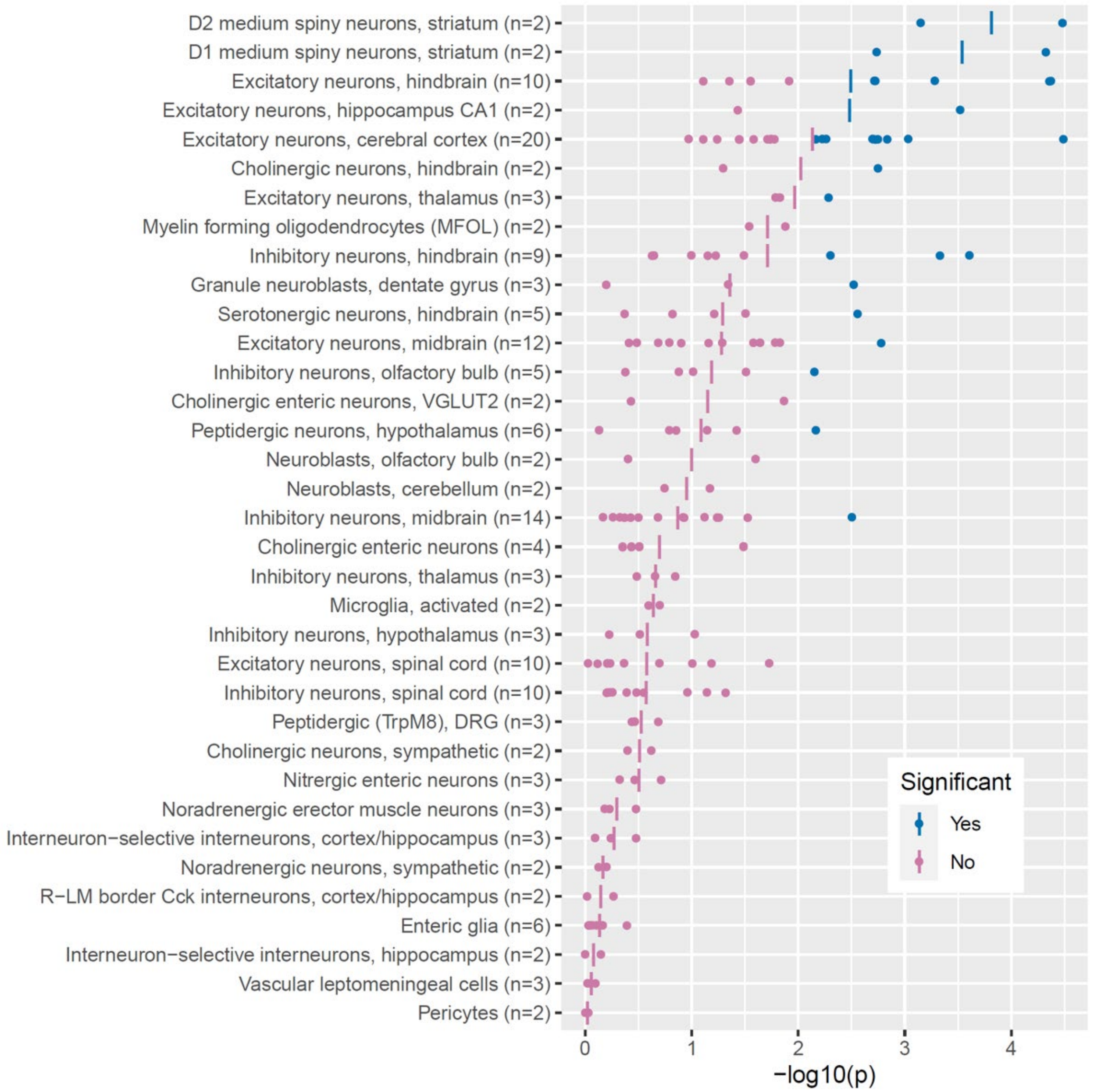


**Supplementary Figure 10**: **Enrichment of TD GWAS signals across 35 cell-type clusters from the mouse brain.** Enrichment analysis was performed using MAGMA, based on gene expression profiles from 265 cell types identified by single-cell RNA sequencing of the whole mouse nervous system[43]. A subset of 166 cell types was grouped into 35 clusters according to their functional and anatomical annotations, with each cluster comprising at least two cell types. Each dot represents the $-\log_{10}(P)$ enrichment value for an individual cell type within its cluster. The vertical bar denotes the mean $-\log_{10}(P)$ across cell types in that cluster. Blue dots and bars indicate clusters or cell types showing significant enrichment of TD genetic signals (FDR < 0.05), while pink dots and bars represent non-significant results.

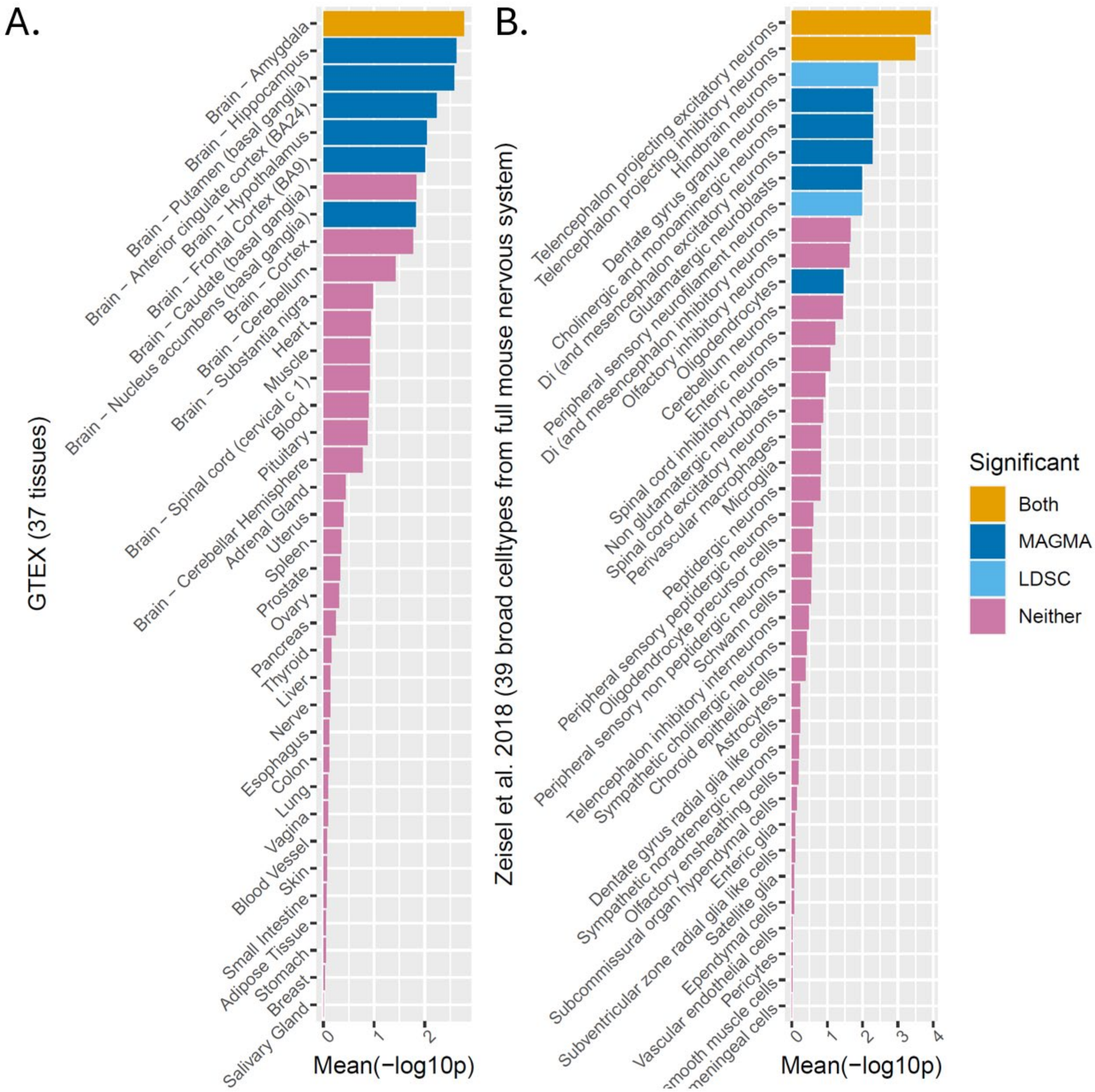


**Supplementary Figure 11. Tissue and cell-type enrichment of TD genetic signals.** A) Tissue enrichment analysis of TD genetic signals across 37 human tissues from GTEx project v8 (ref). B) Cell-type enrichment analysis across 39 broad cell types from the full mouse nervous system, based on single-cell RNA sequencing data from Zeisel et al.[43]. Enrichment analyses were performed using both MAGMA and LDSC. Orange bars indicate tissues or cell types enriched for TD genetic signals in both MAGMA and LDSC (FDR corrected P < 0.05 across 37 human tissues or 39 mouse nervous cell types, respectively). Dark blue bars represent significant enrichment in MAGMA only. Light blue bars represent significant enrichment in LDSC only. Pink bars indicate no significant enrichment in either methods.

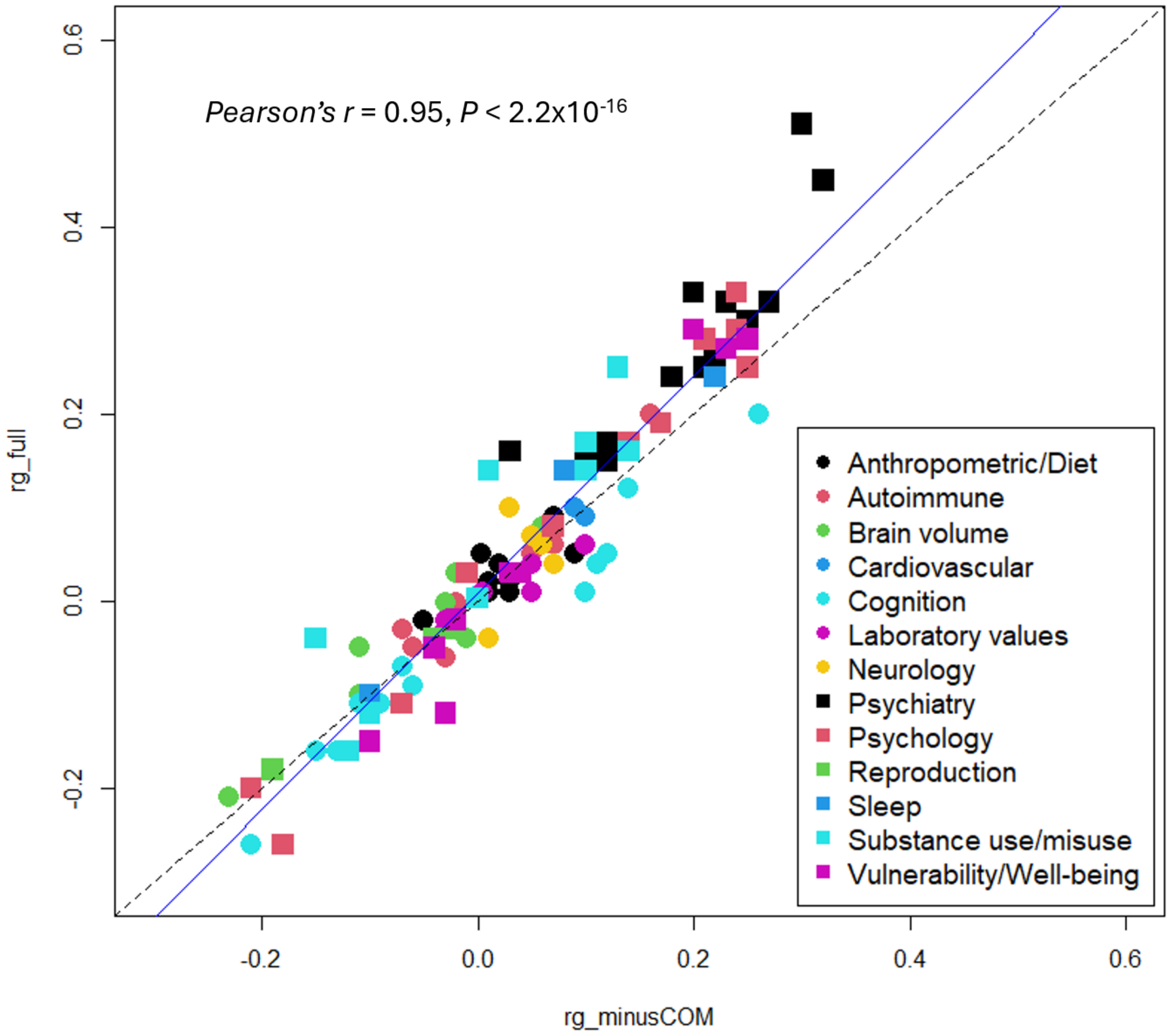


**Supplementary Figure 12. Concordance of genetic correlations with and without cohorts ascertained for other psychiatric disorders.** Scatter plot comparing genetic correlation estimates from the full TD GWAS meta-analysis (rg_full, y-axis) with those obtained after excluding three cohorts ascertained for other psychiatric disorders (rg_minusCOM, x-axis). Each point represents one of the 108 traits analyzed. The black dotted line indicates y = x, representing perfect concordance between estimates. Deviation from this line reflects the impact of excluding comorbid cohorts on $r_g$ estimates. Overall concordance was quantified using Pearson's correlation coefficient (*Pearson's r* = 0.95, $P < 2.2\text{x}10^{-16}$), indicating high consistency between the two sets of estimates. The blue line represents the best-fitting linear regression (β = 1.16; Confidence Interval: 1.09-1.23, $P < 1.9\text{x}10^{-5}$), suggesting inflation of genetic correlation estimates on some traits when including cohorts ascertained for other psychiatric disorders. Traits are color-coded by category as indicated in the legend.
